# Reconciling founder variant multiplicity of HIV-1 infection with the rate of CD4^+^ decline

**DOI:** 10.1101/2024.03.14.24304300

**Authors:** James Baxter, Ch. Julián Villabona Arenas, Robin N. Thompson, Stéphane Hué, Roland R. Regoes, Roger D. Kouyos, Huldrych F. Günthard, Jan Albert, Andrew Leigh Brown, Katherine E. Atkins

**Affiliations:** Usher Institute, Edinburgh Medical School, The University of Edinburgh, Edinburgh, United Kingdom; Department of Infectious Disease Epidemiology, Faculty of Epidemiology and Population Health, London School of Hygiene and Tropical Medicine, London, United Kingdom; Centre for Mathematical Modelling of Infectious Diseases, London School of Hygiene and Tropical Medicine, London, United Kingdom; Mathematical Institute, University of Oxford, Oxford OX2 6GG, United Kingdom; Department of Environmental Systems Science, Institute of Integrative Biology, ETH Zurich, Zurich, Switzerland; Department of Infectious Diseases and Hospital Epidemiology, University Hospital Zurich, Zurich, Switzerland; Institute of Medical Virology, University of Zurich, Zurich, Switzerland; Department of Microbiology, Tumor and Cell Biology, Karolinska Institutet, Stockholm, Sweden; Department of Clinical Microbiology, Karolinska University Hospital, Stockholm, Sweden; Institute of Evolutionary Ecology, The University of Edinburgh, Edinburgh, United Kingdom; *Correspondence to: James Baxter, The University of Edinburgh, Edinburgh, EH9 3FL, United Kingdom

## Abstract

Three quarters of new HIV-1 infections are reported to be initiated by a single genetic variant. Infections initiated by multiple variants have been linked with higher recipient set point viral loads (SpVL) and a faster rate of CD4^+^ T cell decline, indicative of a worse clinical prognosis if left untreated. These findings have not been universally replicated, however, and a mechanism through which multiple variants might lead to a worse prognosis is yet to be elucidated. In this study, we first summarised the existing evidence for this ‘dose response’ phenomenon for HIV-1, and quantified how likely we are to observe a true difference in set point viral load between multiple and single variant infections. Next, we considered whether the association between higher SpVL and multiple variant infection could exist in the absence of a causal mechanism. For a fixed diversity, high transmitter SpVL could simultaneously lead to high recipient SpVL through the inheritance of a ‘high virulence’ genotype and a greater probability that recipient infection is initiated by multiple genetic variants. Nonetheless, a high transmitter SpVL also shortens the duration of infection, consequently reducing the likelihood of the higher SpVL individual transmitting and restricting the overall accumulation of viral diversity. We combined data-driven models of transmission, heritability and HIV-1 disease progression to test whether an association between multiple variant infection and clinical progression is expected. First, we found that we are unlikely to record a significant difference in SpVL between multiple and single variant infections, at frequencies of multiple variant infections consistent with empirical observations. Second, we found that we would not expect multiple variant infections to lead to higher SpVL or faster CD4^+^ T cell decline without a causal mechanism. Specifically, the probability that infection is initiated by multiple variants is greatest at the highest transmitter SpVLs, yet the relationship between transmitter and recipient SpVL is relatively weak. This finding supports the hypothesis that a within-patient causal mechanism is required to explain the association of multiple variant infection with higher viral loads and faster CD4^+^ T cell decline. Further investigation into events happening during and just after transmission are required to enhance our understanding of this association.

## Introduction

Almost all people living with HIV will progress to AIDS in the absence of treatment, and ultimately die of AIDS-related conditions. The progression rate to AIDS, however, varies considerably between individuals ^1,2^. Factors understood to determine the rate of disease progression include characteristics of the infecting virus such as replicative capacity, immunogenicity, or pre-adaptation ^3–5;^ and characteristics of the person such as age and Human Leukocyte Antigen (HLA) phenotype^6–8^.

In some viral infections, a worse clinical prognosis has been associated with an increased inoculum size, often described as a ‘dose-response’ relationship ^9,10^. For HIV-1, infections initiated by multiple genetically distinct viral variants are associated with elevated set point viral loads (SpVL) and faster CD4^+^ T cell decline^11–15^, which are indicative of a faster progression to AIDS when left untreated^3^. Despite 25% of new infections being founded by multiple genetic variants^16–19^, a mechanistic understanding of this dose-response relationship remains elusive^15,20^.

Recipient SpVL is in part explained by viral genotype, which is inherited from the transmitting individual’s infection. As such, transmitting partners with high SpVLs may be associated with recipient infections with high SpVLs ^21–25^.

Concomitantly, the probability of a recipient infection being initiated with multiple variants is greater for an individual with higher SpVL, given identical viral diversity^26^. We might expect, therefore, that we should observe individuals with multiple variant infections to have a higher SpVL because their infection would have been more likely initiated by a ‘high SpVL’ genotype. Nonetheless, high viral loads shorten the duration of infection, thereby reducing the likelihood of the higher SpVL individual transmitting and restricting the accumulation of viral diversity. The impact of SpVL on the dynamics of the transmitter’s infection therefore likely determines the need for a causal mechanism that links multiple variants to increased SpVL.

In this study, we test this assumption by first summarising the existing evidence for an association between infections initiated by multiple variants and faster CD4^+^ T cell decline. We then develop an analytical framework to evaluate how likely a difference in progression to AIDS between individuals with and without infections initiated by multiple variants is to be detected by these observational studies. Finally, we construct a model framework to test the hypothesis that, in the absence of an explicit causal mechanism, we would expect to observe an association between multiple variant infection and prognosis.

## Results

### Observational evidence for an association between infections initiated by multiple variants and faster CD4^+^ decline

We conducted a systematic search of the PUBMED database, which returned 292 unique studies. We identified five studies that characterised associations between founder variant multiplicity, viral load and CD4^+^ T cell decline across six epidemiological cohorts (Table 1)^11–15^. All studies inferred a binary ‘single/multiple’ classification of founder variant multiplicity, and all studies modelled the outcome of multiple variant infection on viral load. Five of the six epidemiological cohorts were analysed for a CD4^+^ T cell outcome. Heterosexual transmissions were analysed in Sagar et al.^11^, Abrahams et al.^12^, and Janes et al.^13^, while men-who-have-sex-with-men (MSM) transmissions were also analysed in Janes et al., Chaillon et al.^14^, and Macharia et al.^15^.

**Table 1:** Summary of the cohort characteristics and key results from studies that have analysed the association between CD4+ cell decline and infections initiated by multiple founder variants. Risk groups: HSX heterosexual, MSM men-who-have-sex-with-men, MF male-to-female. Probability of multiple variants, P, as estimated by the original studies using the methods described. Study design: RCT - Randomised controlled trial. Methods: heteroduplex assays distinguish genetically similar and dissimilar genomic segments using gel electrophoresis of viral RNA; mathematical model and sequence diversity/genetic distance methods test whether the observed diversity fits an assumed threshold or distribution of diversity that would be expected under a hypothesis of neutral exponential growth from a single variant; visual haplotype methods may be used to identify clusters of sequences sharing polymorphisms. Recipient phylogenetic methods test whether the phylogeny estimated from the recipient sequences is consistent with the star-like topology expected for a single variant infection. Phylogenies that use both source and recipient sequences from known transmission pairs allow us to estimate the number of founder variants by counting the number of source phyly nested within the source sequences. Sampling: EDI - estimated date of infection. Analysis: MW Test - Mann-Whitney U Test, LMM - Linear Mixed Model. Effect Size: Sqrt CD4 = Square root CD4+ T Cell Count. Significance: ns; P > 0.05; * P ≤ 0.05; ** P ≤ 0.01; *** P ≤ 0.001 as measured in the original studies.

|  | Number of Individuals | Location | Risk Group | Study Type | Multiple variant detected (%) | Method of variant multiplicity detection | Sampling details | Analysis | Effect Size (with significance) | Analysis | Effect Size (with significance) |
| --- | --- | --- | --- | --- | --- | --- | --- | --- | --- | --- | --- |
| **Sagar 2003** | 156 | Kenya | HSX (MF) | Prospective serodiscordant couples | 57 | Heteroduplex mobility assay | Median 6 samples (range: 1-26) over median 907 (range: 14-2933) days. Median time to first sample: 75 days | M-W Test for primary infection, LMM for longitudinal analysis | + 0.27 (*) | M-W Test for primary infection; Kaplan-Meier curves and log-rank test for longitudinal analysis | (*) |
| **Abrahams 2009** | 90 | South Africa; Malawi | HSX (MF) | Prospective serodiscordant couples (n=69); Observational clinic study acute infections (n = 21). | 22 | Distribution of hamming distances, sequence alignments, recipient phylogenetic topology, mathematical model | 24 individuals with absolute viral load and CD4+ counts at 12 months post-infection | Undescribed univariate test of difference between means | - (ns) | Undescribed univariate test of difference between means; Fisher's Exact Test to compare founder multiplicity and fast/slow progressor status | -(ns) |
| **Janes 2015** | 100 | Thailand (RV144) | HSX | Vaccine RCT | 32 | Sequence alignments, recipient phylogenetic topology and sequence diversity | Initial plasma samples at diagnosis, then 1, 3, 6, 9, 12, 18 and 24 months | Weighted generalised estimating equations | +0.239 (**) | Weighted generalised estimating equations | -2.112Sqrt CD4 (*) |
|  | 63 | Canada; Peru; USA (STEP) | MSM | Vaccine RCT | 25 |  | Initial plasma samples at diagnosis, then at 1, 2, 8, 12, 26, 52, 78 and 104 weeks |  | +0.372 (***) |  | -0.507 Sqrt CD4 (ns) |
| **Chaillon 2016** | 26 | USA | MSM | Case series | 53 | Genetic distance, source/recipient phylogeny topologies | Samples collected mean 70 days (range 11–170) after EDI | Undescribed univariate test of difference between means | - | NA | NA |
| **Macharia 2020** | 38 | Kenya | MSM | Prospective cohort study | 39 | Distribution of hamming distances, sequence alignments, recipient phylogenetic topology. | First sample at 21 (range:12-59) days post-infection. 27 patients on monthly follow-up and 10 on quarterly follow up for 5 years | M-W Test to compare set point viral load (calculation undescribed) | -(ns) | Kaplan-Meier curves and log-rank test | -(***) |

Multiple variant infection was associated with significantly higher viral load at diagnosis in four epidemiological cohorts. To compare viral loads between infections initiated by single and multiple variants, Abrahams et al., Chaillon et al. and Macharia et al., deployed univariate statistical tests. Sagar et al. and Janes et al. implemented linear mixed-effects models and generalised linear models, respectively. Janes et al. included key covariates, such as sex, age and human leukocyte antigen genotype, whereas all other studies reported unadjusted estimates. Effect sizes were reported for three epidemiological cohorts and were broadly consistent (Table 1). For the studies that analysed repeat viral load measurements, the presence of a significant difference between single and multiple variant infections varied throughout infection. For example, at the time of diagnosis, higher viral load was not associated with multiple variant infection in Janes-STEP, but at each time-point thereafter, higher viral load was associated with multiple variant infection. In both epidemiological cohorts analysed by Janes et al, the magnitude of the increase in viral load due to multiple variant infection decreased over time.

Of the four epidemiological cohorts in which a significant association between viral load and multiple variant infection was identified, a further association with lower CD4^+^ T cell count or faster CD4^+^ T cell decline was demonstrated in two. Mean square root CD4^+^ T cell counts over the first year of infection were analysed by Janes et al., revealing a significant decrease in CD4^+^ T cell count associated with multiple variant infection in Janes-RV144. Meanwhile, Sagar et al. had performed a survival analysis of the proportion of individuals with a CD4^+^ T Cell count above 350 cells mm^-3^, stratified by founder variant multiplicity. This analysis revealed women infected with multiple variant infections would reach a CD4^+^ T cell count of less than 350 cells mm^-3^ in 39.2 months (95% Confidence Interval (CI): 30.8 - 47.6), significantly faster than 56.8 months (46.0 - 67.7) for those with single variant infections. A second study, Macharia et al, also identified a significantly greater rate of decline of the proportion of individuals with a CD4^+^ T Cell count above 350 cells mm^-3^, but did not also identify an association with SpVL. One cohort did not demonstrate any association between multiple variant infection and either viral load or CD4^+^ T cell decline^12^.

### Do we expect to observe an association between multiple variant infection and SpVL in an observational study?

Previous observations linking multiple variant infection to a higher SpVL and faster rate of CD4^+^ T cell decline are inconsistent. Therefore, we first tested how likely it is that a true difference in SpVL between individuals with and without infections initiated by multiple variants, would have been detected by the observational studies (Table 1). To compare the probability of observing a significant difference between the log_10_ SpVL of infections initiated by single and multiple variants, we first modelled changes in the distribution of log_10_ SpVL over a transmission cycle. We then approximated the recipient log_10_ SpVL distribution with two Normal distributions of equal variance, representing single and multiple variant infections. The difference in the means was inferred from the signed effect size of observational studies. We then resampled the single and multiple variant log_10_ SpVL distributions proportional to the frequency of multiple variant infection in a population and tested if the samples were significantly different.

Across different study sizes, we found that the probability that a true difference in log_10_ SpVL between individuals with and without infections initiated by multiple variants will be detected by an observational study varies considerably (Figure 1). Given the calculated signed effect sizes of multiple variant infection on log_10_ SpVL by Janes et al. and Sagar et al, and their respective study sizes, we estimated the proportion of observational studies likely to detect a true significant effect of multiple variant infection. For Janes-RV144, we estimated that one would expect to record a significant difference in log_10_ SpVL between single and multiple SpVL in less than half (0.411 (95% CI: 0.381 - 0.441)) of cohorts. Conversely, for Janes-STEP we estimated that one would be equally as likely to record a significant difference than not (0.517 (0.486 - 0.548)), and for Sagar et al. that one would be more likely than not to record a significant difference (0.672 (0.643 - 0.701)).

**Figure 1:**
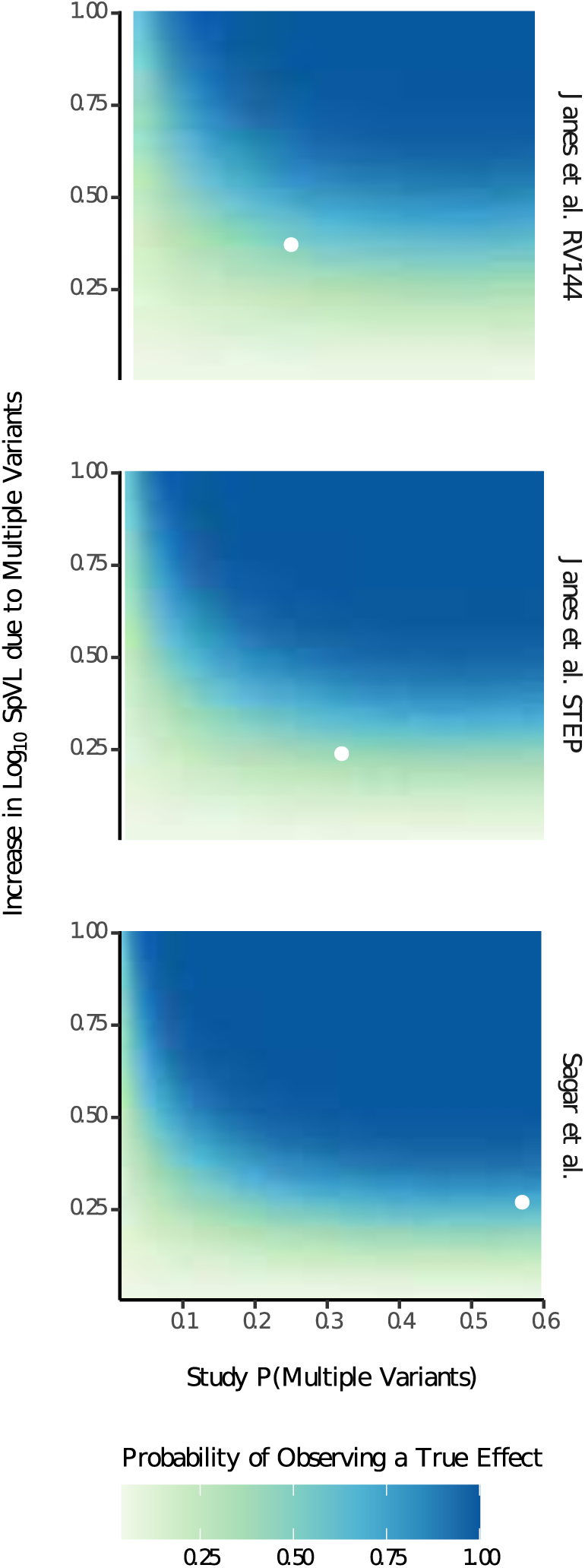
For each study that calculated an effect size, we calculated the probability of observing a true significant difference in SpVL between individuals with and without infections initiated by multiple founder variants. Scaling with study size (n), an effect size threshold is apparent, above which one is more likely than not to observe a significant difference in SpVL between single and multiple variant infections. The probability that infection is initiated by multiple variants greatly increases the probability of observing a true difference in SpVL due to founder variant multiplicity, saturating between 0.2 and 0.3, depending on study size. The effect sizes calculated by each study and the frequency of multiple variant infections are superimposed.

Our analysis also shows that for given effect size, the probability that infection is initiated by multiple variants influences the probability of observing a true difference between multiple and single variant SpVLs. The probability that infection is initiated by multiple variants differs significantly by risk group^19^, therefore analysing one specific risk group could determine how likely one is to record a significant association between multiple variant infection and greater log_10_ SpVL. To illustrate this, we assume a cohort of size 66 and an effect size of 0.27 log_10_ copies ml^-1^ (medians, table 1). The probability of observing a significant difference between the SpVLs of infections initiated by multiple and single variants in a male-to-female cohort is 0.289 (95% CI: 0.262 - 0.318), compared to 0.257 (0.231 - 0.285) for a female-to-male infection cohort, and 0.367 (0.338 - 0.397) for a MSM cohort. Compared to a male-to-female cohort, we are significantly more likely to observe a true difference between the SpVLs of infections initiated by single and multiple in a MSM cohort (Odds Ratio (OR) 1.61 (95% CI: (1.31-1.97)). Conversely, we are significantly less likely to observe a true difference in a female-to-male cohort (OR 0.562 (0.442-0.713)). Overall, we see a lower probability that a true difference in SpVL between individuals with and without infections initiated by multiple variants will be detected by an observational study at probability the infection is initiated by multiple variants less than 0.3. Observational studies are not certain to detect a true difference in SpVL between individuals with and without infections initiated by multiple variants.

### All else being equal, higher recipient SpVLs are associated with a lower probability of multiple variants having initiated infection

To establish whether we should expect to observe a significant association between the probability that infection is initiated by multiple variants and two markers of HIV prognosis, the SpVL and the rate of CD4+ T cell decline, in the absence of an explicit causal mechanism, we combined three models characterising HIV-1 transmission and disease progression (Figure 2a). First, we calculated the probability that a recipient’s infection was initiated by multiple variants as a function of the transmission pair’s SpVL (the ’transmission model’). Second, we calculated the recipient partner’s SpVL as a function of their transmitting partner’s SpVL (the ’heritability model’) (Figure 2b). Third, we calculated the rate of a recipient’s CD4 cell decline as a function of their SpVL (the ’tolerance model’) (Figure 2c)^27^. Using these models, we were able to infer the extent to which SpVL or CD4^+^ T cell decline varied as a function of the probability that infection was initiated by multiple variants in a risk-group stratified population of people living with HIV. Importantly, our model assumes no causal relationship between multiple variant infection, higher SpVL or the rate CD4^+^ T cell decline.

**Figure 2:**
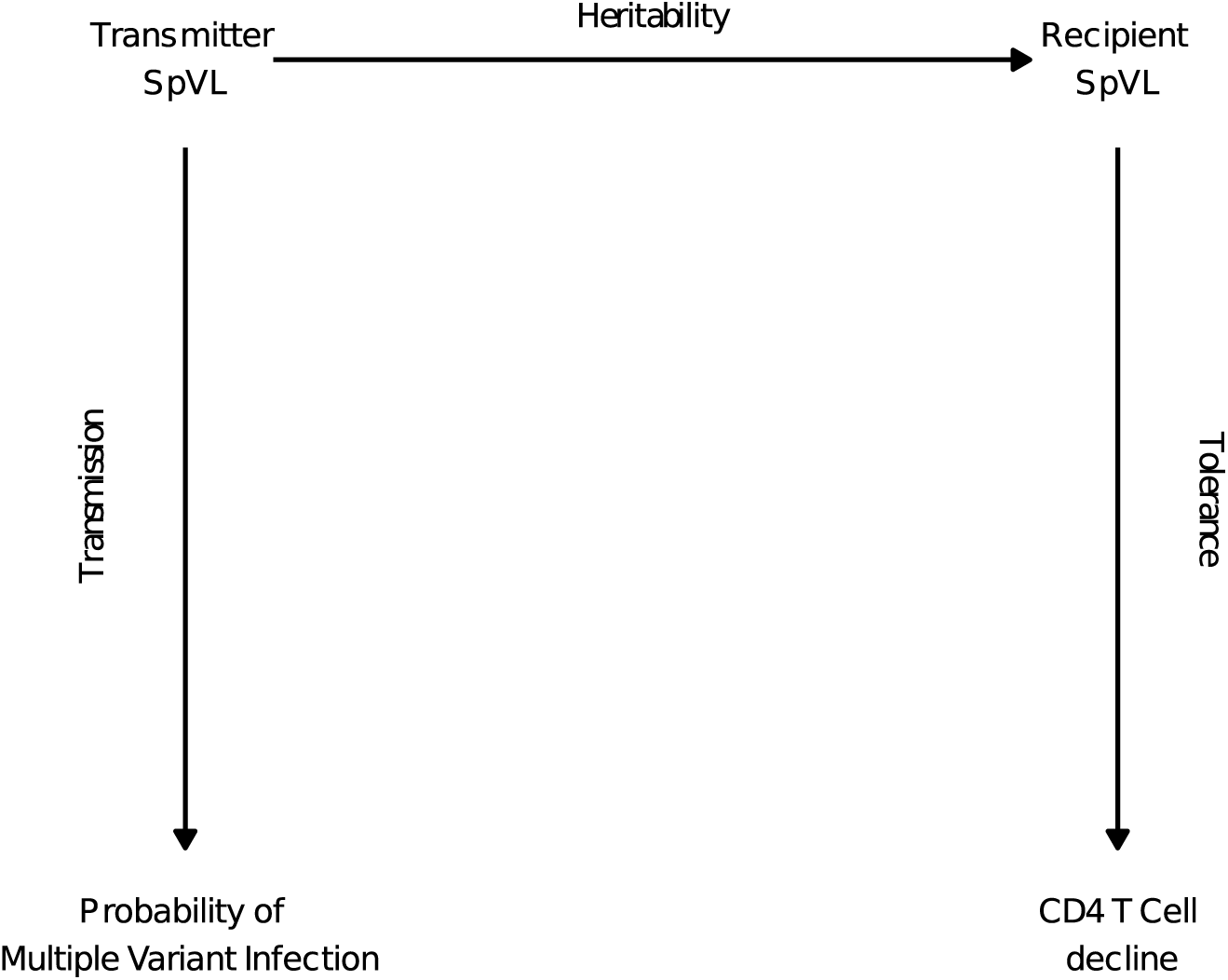
A) To evaluate the relationship between the probability that infection is initiated by multiple variants, and the rate of CD4+ T cell decline, we leverage three well-characterised empirical relationships: we predict recipient set point viral load as a function of transmitter SpVL; calculate the probability that a potential recipient infection is initiated by multiple variants, as a function of transmitter SpVL; and predict the daily rate of CD4^+^ T cell decline (ΔCD4) from recipient SpVL.

**Figure 2:**
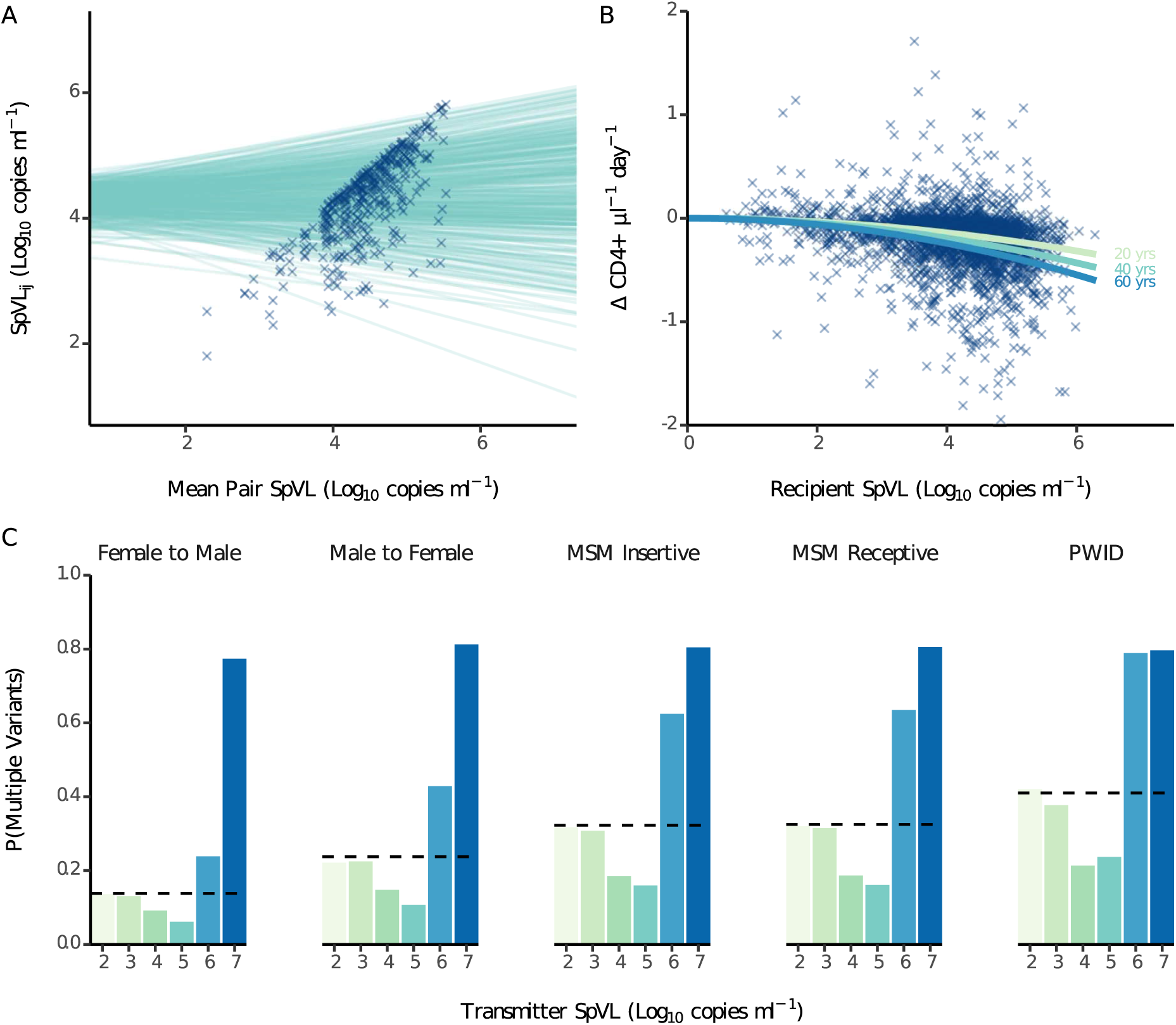
Component models of the analytical framework outlined in figure 2: A) Heritability model: A linear mixed model predicting the SpVL for individual j in transmitter-recipient pair i as a function of the pair’s mean SpVL; B) Tolerance model: a non-linear model that predicts the rate of CD4^+^ T cell decline for a given recipient SpVL, accounting for linear effects of age and sex. C) Transmission model: A mechanistic model to calculate the probability that an infection is initiated by multiple variants, given a transmitter SpVL. We fitted this model for each risk group using previously estimated probabilities of acquisition and that infection was initiated by multiple variants (dashed line).

The heritability and transmission models were newly fitted for this study. We estimated the broad sense heritability, *H*^2^, of log^10^ SpVL using a Bayesian linear mixed model and calculated an approximate R^2^ value. Assuming a random intercept for each pair, and adjusting for recipient sex, age at infection, partner, and risk group, we estimated the *H*^2^ to be 0.238 (95% Highest Posterior Density (HPD): 0.123-0.341). When fitting our transmission model, the probability that infection is initiated by multiple variants was lowest at intermediate transmitter SpVLs (P(MV) = 0.14 at 4 log_10_ copies mm-3 for male-to-female transmissions), but more likely at higher transmitter SpVLs (P(MV) = 0.81 at 7 log_10_ copies mm-3 for male-to-female transmissions) (Figure 2D). Lower transmitter SpVLs were approximately representative of the risk group average probability that infection was initiated by multiple variants (P(MV) = 0.22 at 2 log_10_ copies mm-3 for male-to-female transmissions).

Applying our model framework to simulated risk-group stratified transmission pairs, we inferred relationships between the probability that infection is initiated by multiple variants, recipient SpVL, and the rate of CD4^+^ decline. Across risk groups, we found that recipient infections were most likely to have a low probability of being initiated by multiple variants and present with intermediate log10 SpVL (Figure 3a). Specifically, the median probabilities that infection was initiated by multiple variants were 0.173 (IQR: 0.147-0.221) for men-who-have-sex-with-men:receptive (MSM-receptive), 0.212 (0.178-0.276) for people-who-inject-drugs (PWID), 0.131 (0.108-0.167) for male-to-female, 0.080 (0.064-0.104) for female-to-male, and 0.174 (0.146-0.222) for men-who-have-sex-with-men:insertive (MSM-insertive). For risk groups associated with a greater risk of multiple variant infection, such as MSM-receptive and PWID, most infections would be expected to have a low probability of being initiated by multiple variants, except a small proportion of infections initiated with multiple variants with very high probability (∼0.8). There was no trend between these relatively high probabilities that infection is initiated by multiple variants and high recipient log10 SpVL.

**Figure 3:**
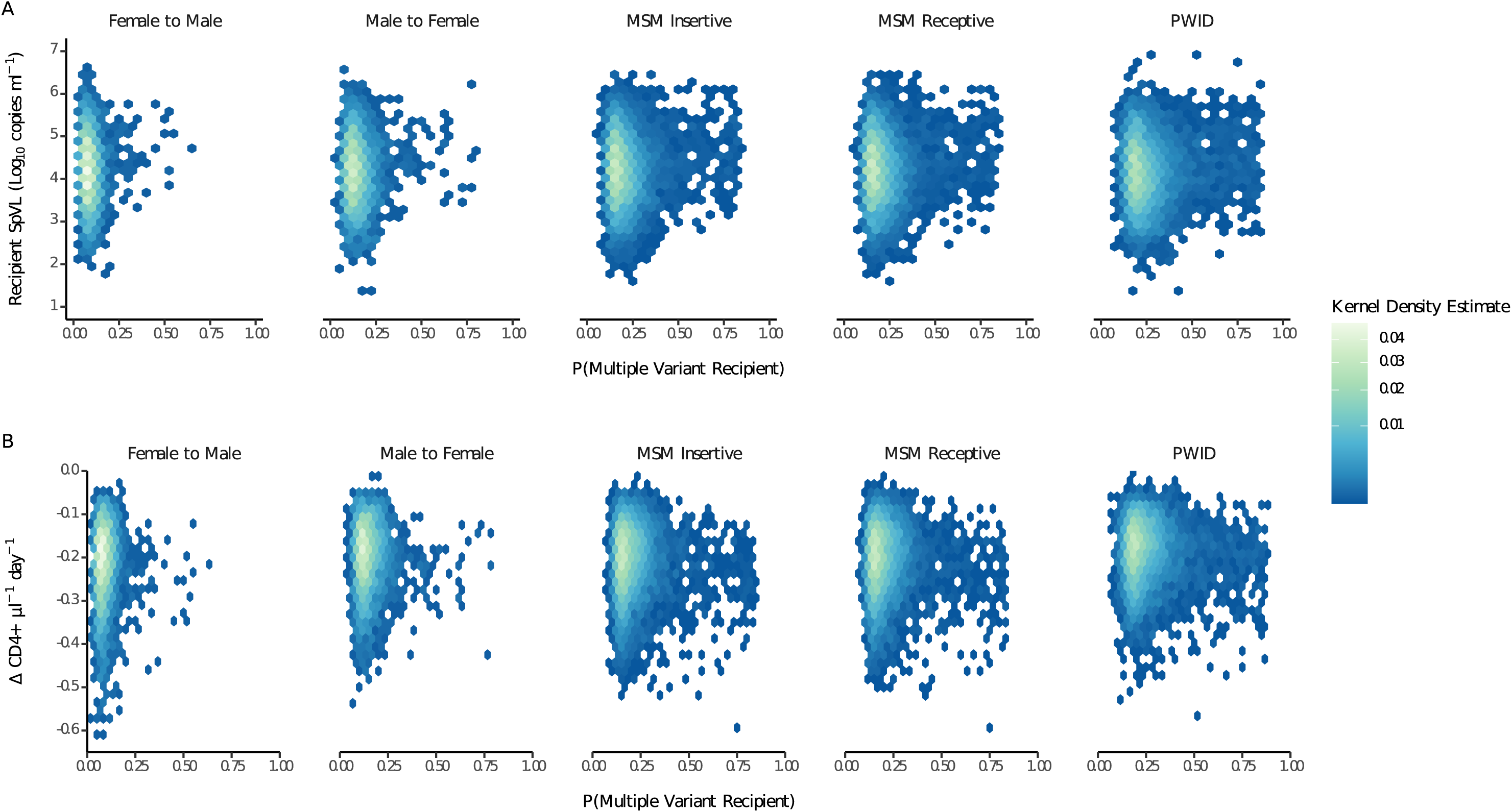
Kernel density estimates of recipient SpVL and the probability that the infection was initiated by multiple variants (A), and the rate of decline of CD4^+^ T cells and probability that infection was initiated by multiple variants (B), stratified by risk group.

Across risk groups, we predicted a relatively slow rate of CD4^+^ T cell decline, determined as a function of recipient log10 SpVL, age and sex. Specifically, the median rates of daily CD4^+^ T cell decline were -0.200 (IQR: -0.250- - 0.154) for MSM-receptive, -0.177 (-0.225- -0.138) for PWID, -0.189 (-0.239- -0.145) for male-to-female, -0.204 (-0.260- -0.158) for female-to-male, and -0.200 (-0.250- -0.154) for MSM-insertive (Figure 3b). For risk groups associated with a greater probability of multiple variant infection, such as MSM-receptive and PWID, the daily rate of CD4^+^ T cell decline remains consistent with risk groups associated with a lower probability of multiple variant infection. Where an infection was predicted to be initiated with multiple variants at a high probability, this did not correspond to a faster rate of CD4^+^ T cell decline. Qualitatively, our findings were consistent across transmitter allocation methods and when SpVL and CD4^+^ T cell decline were compared with the probability that infection is initiated by multiple particles (Figures S10 & S11, pages 18-19).

To compare our model framework to the outcome variables used in observational studies, we calculated the mean SpVLs for single and multiple variant infection as well as the proportion of each risk group that would have an excess of 350 CD4^+^ T cells per mm^3^ over time. The average SpVL of female-to-male infections initiated by multiple variants was the same as single variant infections (median 4.22 (IQR: 4.20-4.25) log_10_ copies mm^-3^; 4.22 (4.22-4.23) log_10_ copies mm^-3^, respectively). Conversely, the average SpVLs of male-to-female, MSM-receptive, MSM-insertive and PWID infections initiated by multiple variants were slightly greater than infections initiated by a single variant (4.18 (4.16-4.20) log_10_ copies mm^-3^ and 4.17 (4.16-4.17) log_10_ copies mm^-3^; 4.24 (4.22-4.25) log_10_ copies mm^-3^ and 4.23 (4.22-4.23) log_10_ copies mm^-3^; 4.24 (4.22-4.25) log_10_ copies mm^-3^ and 4.23 (4.22-4.23) log_10_ copies mm^-3^; 4.17 (4.16-4.18) log_10_ copies mm^-3^ and 4.15 (4.15-4.16) log_10_ copies mm^-3^ respectively). Across all risk groups, log_10_ SpVLs of single variant infection were significantly less dispersed than those from infections initiated by multiple variants (paired, single tailed t-test, p = 0.0027).

We also found that the difference in time to a clinically relevant (≤ 350 cells mm-3) CD4^+^ T cell count between multiple and single variant infections varied slightly across risk groups (Figure 4b). In MSM-receptive transmissions, the time to a clinically relevant CD4^+^ T cell count for single variant infections was 9.96 years, compared to 9.92 years for infections initiated by multiple variants. In both male-to-female and PWID transmissions, however, the median time to a clinically significant CD4^+^ T cell count in multiple variant infections was the same as that for single variant infections (10.7 and 10.9 years, respectively). In female-to-male and MSM-insertive transmissions, the time to a clinically significant recipient CD4^+^ T cell count for infections initiated by a single variant (9.78, 9.95) was actually slightly shorter than those initiated by multiple variants (9.93, 9.97). Across all risk groups, the median time to a clinically significant recipient CD4^+^ T cell count associated with single variant infection were significantly less dispersed than those from infections initiated by multiple variants (paired, single tailed t-test, p = 0.003).

**Figure 4:**
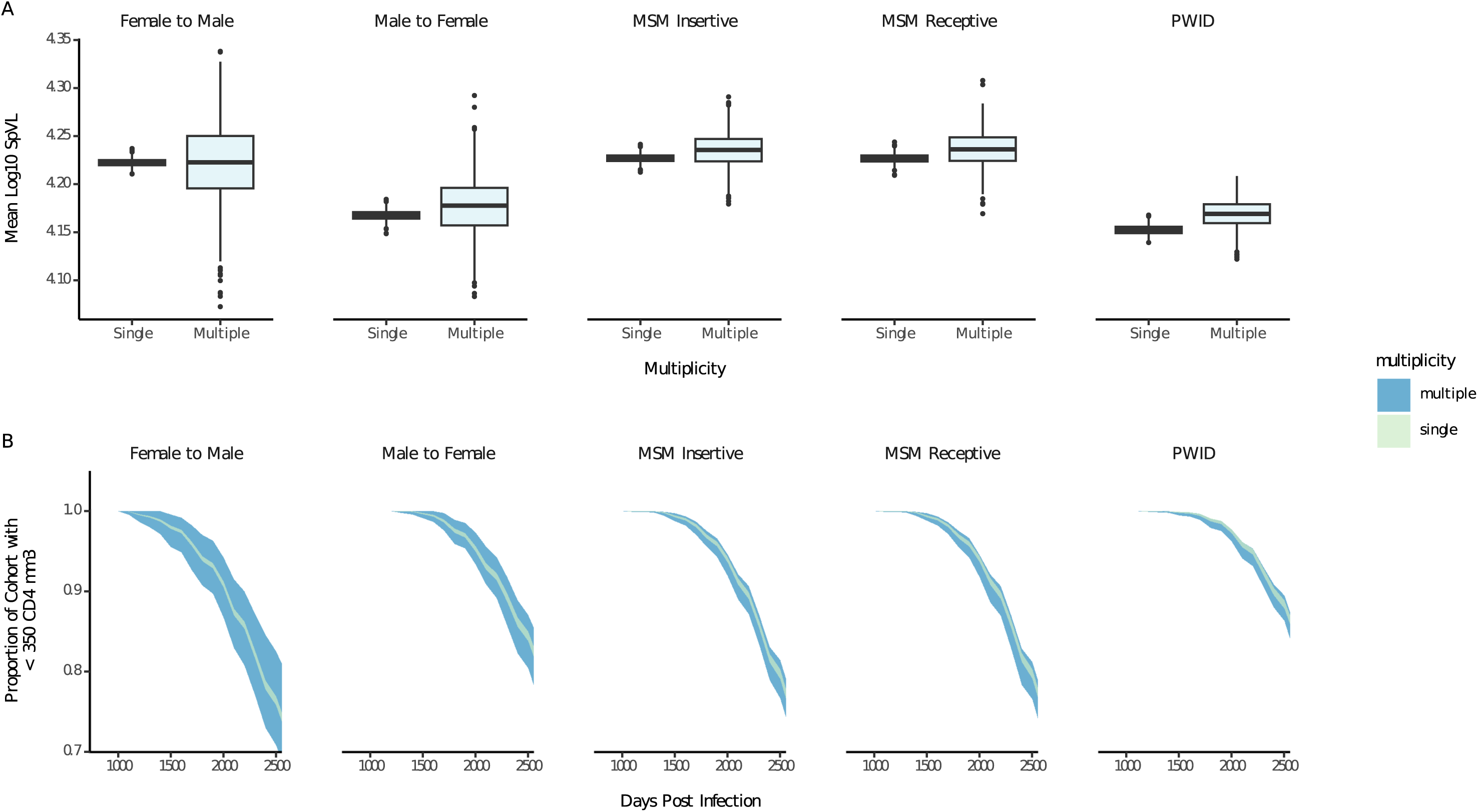
A) Box plots comparing the distribution of the re-sampled mean set point viral loads across transmission groups. B) extrapolating the proportion of individuals with less than 350 CD4 T cells/mm^3^ from linear rates of decline and resampling according to the probability that infection was initiated by multiple variants. Upper and lower bounds of the ribbons represent the 5th and 95th quantiles.

## Discussion

In this study, we sought to understand how likely we are to observe a significant association between infections initiated by multiple variants and higher SpVL or faster CD4^+^ T cell decline. First, we identified six epidemiological cohorts in which associations between founder variant multiplicity, viral load and CD4^+^ T cell decline had previously been investigated. We then showed how the probability of observing a true difference in SpVL due to multiple variant infection might depend on the study parameters and the risk group analysed. We tested whether we would expect to observe an association between multiple variant infection and prognosis in the absence of an explicit causal mechanism. We identified non-linear relationships between the probability that infection is initiated by multiple variants and SpVL or CD4^+^ T cell decline. Nonetheless, log10 SpVL and the rate of CD4^+^ T cell decline were generally lower at higher probabilities of multiple variants. Across risk groups, multiplicity-stratified predictions of log10 SpVL differed slightly. Specifically, log_10_ SpVLs of male-to-female, MSM-receptive, MSM-insertive and PWID infections initiated by multiple variants were slightly greater than infections initiated by a single variant.

Both log10 SpVL and the time to a clinically significant CD4^+^ T cell count, were significantly greater variation in multiple variant infections than infections initiated by a single variant across risk groups.

Altogether, our results are consistent with the hypothesis that a causal mechanism is required to recapitulate the observed association between multiple variant infection and HIV-1 prognosis. This suggests that a simple association between higher viral load and greater diversity in the transmitter that leads to higher viral load in the recipient, irrespective of the number of variants initiating infection, is itself an insufficient explanation. Across an infection cycle, individuals with intermediate SpVL infections are more likely to pass on their infection than those with higher viral loads^28^. According to our transmission model infections acquired from such individuals are initiated by multiple variants with low probability while multiple variant infections initiated by individuals with high SpVL are much more likely. Accordingly, to observe an association between high SpVL and multiple variant infection without a within-patient mechanism, SpVL would need to be predominantly determined by the transmitted genotype. Instead, we calculated the broad sense heritability of log_10_ SpVL to be 23.9%, meaning the SpVl of the recipient infection is only weakly determined by the genotypic load of the transmitter virus population. Therefore, a high SpVL genotype is no more or less likely to initiate a high SpVL infections than low SpVL genotype.

Presently, a causal link between the number of variants initiating HIV-1 infection and disease prognosis is yet to be identified. Where an infection is initiated by multiple variants, this is associated with a weaker selective transmission bottleneck such that any ‘reasonably fit’ virus may initiate infection^29,30^. Founder viruses isolated from patients whose infection was initiated by multiple variants have shown a worse replicative capacity in vitro than those initiated by single variants^15^. This may suggest it is not an inherent virological property of the founder viruses that drives an association with a faster rate of CD4^+^ T cell decline, but instead how the presence of multiple variants determines disease dynamics in early infection. Multiple variant infections typically possess one major variant and one or more minor variants at a single point in time^15,31,32^. The relative proportions of these variants in major compartments have been shown to vary throughout early infection, in addition to prolific inter-variant recombination^31,33,34^. Nonetheless, key phenotypes such as coreceptor usage efficiency, IFN-*α* resistance, and viral fitness are not associated with proportions of founder variants in each individual during acute infection^20^. It has been speculated that the shifts in variant proportion in early infection are immune-driven, with minor variants cyclically out-competing the dominant variant due to a non-negligible selective advantage^31^. Given that multiple variant infections lead to earlier presence of broadly neutralising antibodies, it may follow in trying to mount a sufficiently broad immune response, initial efficacy is attenuated and leads to increased sequestration and higher viral loads during the acute phase^35,36^.

We were unable to account for several contributors to HIV-1 disease progression in our models. The role of human genetic variability, in particular specific HLA genotypes that modulate resistance to infection is well documented, and may have a key role in determining SpVL, in particular when transmitter and recipient are phenotypically similar^8,37^. Crucially, the interaction HLA and viral genotype also determines SpVL in the recipient, contravening assumptions of independent action in our basic model of heritability^5^. Viral properties, such as immunogenicity and co-receptor usage, may also contribute to the rate of CD4^+^ T cell decline independent of SpVL, but remain unaccounted for^38^. Our model framework also highlights limitations in our understanding of HIV-1 transmission. Our transmission model assumes no mutation, selection and recombination; all of which may influence the within-person fitness of the infecting virus^33,34^. The heritability model links the within-person processes of the transmission model to the tolerance model; mid-parent offspring regression estimates of heritability are infamously low-powered, however, and we cannot exclude the possibility that within phylogenetically linked transmission pairs, infection of a seronegative partner occurred from an unknown third party or a common source^39^. Furthermore, the precise quantification of pathogen trait heritability remains challenging, with estimates of HIV-1 virulence heritability varying between 7% and 40%^21–25,40,41^.

Our model results indicate that we would not generally expect to observe an association between multiple variant infection and HIV-1 disease progression in the absence of a causal mechanism. Moreover, even given the existence of a true effect, we are not certain to detect it in observational studies. Ultimately, there remain key shortcomings in our existing knowledge of events happening both during and just after transmission that require further investigation to enhance our understanding of this phenomenon.

## Methods

### Literature Review

We first conducted a review to understand the extent to which a difference in HIV-1 prognosis between infections initiated by single and multiple variants is supported. To systematically collate these data, we searched the PUBMED database with the following search criteria: (“HIV”[Title] OR “human immunodeficiency virus type 1”[Title]) AND (“founder*”[Title] OR “multiplicity”[Title] OR “variant*”[Title]) AND (“viral loads”[Title/Abstract] OR “CD4”[Title/Abstract] OR “disease progression”[Title/Abstract] OR “poisson”[Title/Abstract])). Our search was restricted to studies published between 1st January 2000 and 1st September 2023. Briefly, our search strategy required studies to have reported original estimates of founder variant multiplicity in people with acute or early HIV-1 infections; the recipient partner should not be receiving pre-exposure prophylaxis, and the transmitting partner should be antiretroviral treatment naive. No restrictions were placed on study design, geographical location, or age of participants. We then identified a subset of studies that satisfied our search criteria and conducted statistical analyses comparing clinical biomarkers of disease progression between single and multiple variant infections of people living with HIV-1. We recorded the number of study participants, the location of the study, the proportion of the study whose infection was initiated by multiple variants, the method of founder variant quantification, the sampling strategy of viral load and CD4+ measurements, and details of the statistical analysis conducted and any covariates included.

### Probability of observing a significant association between SpVL and Multiple Variant Infection

We extended a previously published framework to describe the distribution of SpVL in the HIV population^42^. In the original framework, the population is subdivided into carriers, representing people living with HIV prior regardless whether they will go on to be transmitters; transmitters, representing carriers that have been selected for transmission; and recipients, representing individuals with recently initiated infections from the transmitters. Making the simplifying assumption that the population of log_10_ SpVL in each subdivision follows a Normal distribution (Supplementary material; Figure S1), the change in mean and variance of each may be expressed as a series of linear equations. We parameterised the carrier population with mean 4.74 log_10_ copies ml-1 and variance 0.78 ^28^.

We then extended this framework to consider the findings that infections initiated by multiple founder variants are associated with significantly higher SpVLs than those initiated by single variants. We assume that the Normal distribution of recipient SpVLs, *X*, calculated by Bonhoeffer et al.’s model, with mean, *μ*, and variance, *σ*^2^, can be approximated by a mixture distribution of two Normal distributions (Supplementary material; Figure S1, page S6). These respective distributions describe the population of SpVLs attributable to multiple and single variant infections in the transmitter population, contributing to the mixture according to their frequency, *p* and (1 − *p*), respectively. The mixture distribution is parameterised by ^45^:

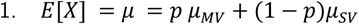

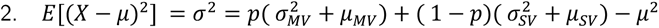

Where 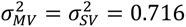^13^, a known effect size representing the true difference in SpVL, *μ_MV_* - *μ_SV_*, and known values of *μ* and *p*, the component distributions may be calculated. We inferred component distributions for all combinations of effect size between +0.01 and 1 log_10_ increase SpVL, and values of p, between 0 and 0.6. We sampled from the component distributions proportionally to the probability that infection was initiated by multiple variants, *p*, and inferred the significance of the difference between the sample means using a one-sided t-test. The proportion of samples with a significant difference between single and multiple variant distributions from 100 independent draws was recorded.

### A model framework to test whether we should expect multiple variant infections to associate with higher viral loads and faster CD4^+^ T cell decline

#### Heritability Model

A proportion of variation in set point viral load between individuals within a putative transmission pair can be explained by genetic variation of the transmitted virus^21,23^. To predict log_10_ SpVL, 6, for individual 7 in pair 8, we fitted a Bayesian linear mixed model to SpVLs of 196 previously determined transmission pairs from the Swiss HIV Cohort Study (SHCS)^43,44^. SpVL was defined as the geometric mean of viral loads over a period of at least 180 days, obtained prior to commencing antiretroviral treatment and excluding the primary (90 days post estimated date of infection) and late (CD4^+^ T cell count was below 100 per µl) phases of the infection. For each pair, we fitted a random intercept, *γ*_0*i*_, set by the mean pair log_10_ SpVL, and adjusted for key covariates: recipient sex (male, female), age at infection (categorised to intervals of: 16-24, 25-29, 30-39-40-80), partner (transmitter/recipient), and risk group (male-female, female-male, men-who-have-sex-with-men).

As the direction of transmission is not known for these pairs, for the purpose of our modelling scenario, we imputed an epidemiological role (transmitter or recipient) for each individual. First, we calculated a theoretical distribution of transmitter log_10_ SpVLs in a population, using a linear equation framework described by Bonhoeffer et al.^42^. We then assigned the role of transmitter within each pair to the individual whose log_10_ SpVL had the highest likelihood of being a transmitter. We conducted two sensitivity analyses on the allocation of transmitter; i) refitting the heritability model with randomly assigned transmitter status within a transmission pair and ii) assigning the individual with highest viral load as the transmitter.

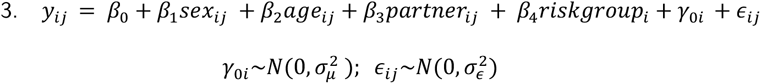

Priors for the parameter values were informed by Hollingsworth et al. (Table S1, page S9)^21^. The model was fitted using Markov Chain Monte Carlo (MCMC) simulation for 10000 iterations across 4 chains. The first 1000 iterations of each chain were discarded as burn-in. Convergence of the joint posterior distribution and adequate mixing of all chains was assessed against criteria of Effective Sample Size, ESS > 1000, and rank-normalised Gelman and Rubin statistic, *R̂*<1.05^45^. Assumptions of normality and homogeneity of variance were satisfied following visual inspection of QQ and residual plots (Figure S4&S5, pages S10-S11) (Table S2, page S12). Predicted values of heritability were drawn from the posterior predictive distribution, calculated as the variance of the predicted SpVL divided by the sum of the variance of predicted SpVL and the expected variance of the errors (Figure S6, page S13)^46^.

#### Tolerance Model

We used a previously described statistical model that characterises the relationship between recipient SpVL and CD4^+^ T cell decline has been described previously^27,47^. In brief, this model was fitted to SpVLs and longitudinal CD4^+^ T cell counts of 3036 people living with HIV-1 who participated in the SHCS. In their analysis, Regoes et al. found that the rate of change of CD4^+^ T cells per ml of blood per day, y, for recipient, j, was best expressed as a quadratic function of SpVL, log_10_ *V_j_*. The tolerance at birth for females is given by, α_(0,F)_, which thereafter changes linearly with age, a, at rate c*_F_*, per year. The sex difference of tolerance at birth, η_(0,M)_, and sex difference between the change of tolerance per life year, M_(0,M)_, represent the linear effects of recipient age and sex, and their interaction.

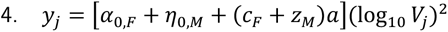

During model fitting, Regoes et al. excluded the linear viral load term and the intercept, finding that they did not deviate significantly from zero. The parameter values used in this study are detailed in the supplementary material (Table S3, page S14).

#### Transmission Model

We extended a previously described transmission model that links transmitter infection viral load with the probability of recipient infection being initiated by multiple viral variants^26^. The model calculates the number of virus particles, *n*, and number of viral variants, N, initiating infection in the recipient partner at a given time *τ* during the transmitter partner’s infection. The number of virus particles founding infection at transmission time, *τ*, is distributed binomially, *n*(*τ*)∼*Bin*(*v*(*τ*),*p*), assuming an inoculum of size, *v*(*τ*), which is proportional to viral load at transmission and a per-particle establishment probability, *p*. The marginal probability that a given number of variants initiates infection, *P*(*N*(*τ*)), is given by 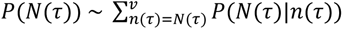. As in the original model, we reconciled the low probability of infection acquisition with a relatively high proportion of multiple variant infections, by assuming only a fraction, *f*, of exposures occur in an environment conducive to transmission resulting in a zero-inflated probability distribution.

The number and diversity of virus particles are modelled across the duration of the transmitter infection. Similar to Thompson et al., we assumed that no transmission occurs during AIDS due to the severity of illness and categorised the remaining infectious period into primary, chronic and pre-AIDS. We then extended the model to relax the previous assumption of fixed viral loads in primary and pre-AIDS infection (Figure S7A, page S15). Specifically, we assumed that viral load varied by Fiebig Stage during primary infection, with stages of length 5 days (stage I), 5.3 days (stage II), 3.2 days (stage III), 5.6 days (stage IV), and V 69.5 (stage V) days^48^. Within each stage, we assumed the log10 viral load profiles follow a normal distribution, and calculated the probability that infection was initiated by multiple variants for the 0.1, 0.15, 0.5, 0.85, 0.9 percentile viral load, weighted by their respective probability density (Figure S7B, page S15). For pre-AIDS infection we fitted a lognormal distribution to previously described viral loads using maximum likelihood, then calculated the probability that infection was initiated by multiple variants at discrete intervals, weighted according to their respective probability density^49^. The best-fit distribution for pre-AIDS viral loads was parameterised by mean log 1.62 and standard deviation log 0.114 (Figure S7C, page S15). Finally, we followed Thompson et al. in assuming that a small number of variants predominate early infection, giving way to a more uniform distribution of variants (synonymous with higher diversity) as infection progresses (Figure S7E, page S15)^50^. We modelled the proportion of the x^th^ most common viral variant at time, [, since infection, as a gamma distributed random variable, X∼*Γ*(*j*, *k*), with shape parameter, *j* = 0.417, and scale parameter, *k* = *τ*/0.563. All other fixed parameters remained the same as first presented by Thompson et al. (Table S4, page S14).

To calculate the probability that an infection is initiated by multiple variants for a given transmitter SpVL, we first estimated risk group stratified parameter values f and *p,* integrated across all infected potential transmitters in the population, and all times during their courses of infection. Whereas Thompson et al. set the per particle transmission probability, p, and the proportion of the time the recipient environment is conducive for transmission, f, such that the conditional probability of transmitting multiple variants in a single act was 0.3, we used a Bayesian model fitting approach to infer joint posterior distributions of *f* and *p* (Figure S9, page S16). We ran two independent MCMC simulations for 30000 iterations with a 25% burn-in for each risk group. After concatenating the two chains, a minimum ESS of 2000 was achieved across all analyses, with a Gelman-Rubin statistic of *R̂* <1.05. Joint posterior distributions of *f* and *p* recapitulated empirical distributions of the per-event probability of acquisition and the probability that infection is initiated by multiple founder variants (Figure S10, page S17)^19,51^.

Parameter values of *f* and *p* were drawn from the joint posterior distribution and used to calculate individual probabilities that an infection is initiated by multiple variants, for a given transmitter SpVLs integrated over the course of that transmitter’s infection.

### Risk-group stratified cohorts

To disentangle variation associated with sex and age across risk groups, we simulated risk group-stratified data from the 196 SHCS transmission pairs used in our heritability analysis. First, we selected only those pairs whose risk group were concordant, and calculated separate variance-covariance matrices for each risk group of mean log10 SpVL, age at infection and sex-pairs. Next, we used these relationships to synthesise populations of size 200, from a truncated multivariate normal distribution. Categorical variables were enumerated prior to simulation and re-classified according to the cumulative probability distribution calculated for each variable from the empirical data. Truncation was applied only to ‘age at infection’ with a lower bound of 16. We validated these results by comparing the difference in the means and variance between the stratified simulated and empirical data.

## Computation

All analyses were implemented in R v4.1.2^52^. The heritability model was fitted in Stan using BRMS v2.20.4^53,54^. Extensive use was made of the Tidyverse suite v2.0.0 for data handling; and tidybayes v3.0.6, emmeans v1.8.9 , bayesplot v1.10.0 and performance v0.10.5 for post-processing^55–58^.

## Ethics

The tolerance and heritability models were fitted to data from the Swiss HIV Cohort Study (SHCS)^44^. The SHCS has been approved by ethics committees of all participating Swiss institutions. Data collection was anonymous and written informed consent was obtained from all participants.

## Acknowledgements

The authors would like to express their gratitude to the participants of the Swiss HIV Cohort Study and to Emma Pujol Hodge and Dr Hannah Lepper for helpful discussions.

## Authors’ Contributions

KEA conceptualised the study. JB and KEA designed the study. RR, RK and HG collected and supplied the data. JB and CJVA analysed the data. JB drafted the manuscript, with discussions and critical revisions from all authors. KEA, ALB, JA and SH supervised the study. All authors had final responsibility for the decision to submit for publication.

## Data Availability

The data used to fit the heritability model from the Swiss HIV Cohort Study are available from the corresponding author on reasonable request.

## Code Availability

All code is available on GitHub under GPL-3.0 licence (https://github.com/J-Baxter/foundervariantsHIV_prognosismodelling).

## Supplementary Methods

### Probability of observing an association between SpVL and Multiple Variant Infection

To decompose the recipient log_10_ SpVL population to a mixture of two weighted Normal distributions corresponding to single and multiple variant infection, we first followed a series of previously described linear equations ^42^. In their study, the distribution of SpVLs were approximated by a Normal distribution, from which the change in the population of SpVLs across a transmission cycle was described. This relies on the concept that the convolution of two Normal distributions is itself a normal distribution, with mean and variance:

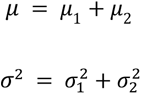

The SpVL phenotype in a carrier population follows the distribution *N*(*μ_cp_*, *σ_cp_*), and comprises the sum of genotypic and environmental effects. Both factors are assumed to be independent and follow Normal distributions *N*(*μ_cg_*, *σ_cg_*) and *N*(*μ_ce_*, *σ_ce_*), respectively:

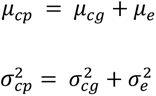

We assume that the population of interest is at equilibrium, allowing us to derive a value of 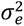 for a given heritability. We assume that broad sense heritability, *h*^2^, of SpVL is 0.34^42^.

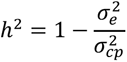

The transmission potential describes the expected number of transmissions deriving from one index individual as a function of set point viral load, integrated over that individual’s asymptomatic infection ^28^. Here, we approximate this quantity to follow a Normal distribution with mean, *μ_o_* = 4 and standard deviation, *σ_o_* = 1 ^42^. Applying the transmission potential to the genotypic and phenotypic mean and variance of the carrier population, the transmitter population with genotypic mean and variance, 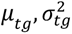 and phenotypic mean and variance 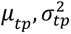 may be calculated:

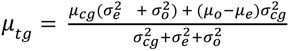

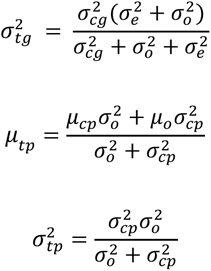

The population of recipient genotypes is assumed to follow the same distribution as transmitter phenotypes with mean, *μ_rg_*, and variance, 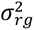:

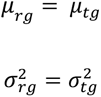

The population of recipient phenotypes is calculated from the genotypic mean and variance, and environmental mean and variance:

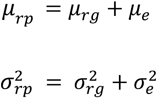

### Mixture distribution of single and multiple variant SpVLs

To compare the probability of observing a significant difference between the log10 SpVL of infections initiated by single and multiple variants, we first modelled changes in the distribution of log10 SpVL over a transmission cycle. We then approximated the recipient log10 SpVL distribution with two Normal distributions of equal variance, representing single and multiple variant infections. However, it is not mathematically exact that the overall recipient distribution should remain Normal when comprised of two weighted components of equal variance. To empirically justify this assumption, we sampled from each component distribution to replicate a recipient population with 25% multiple variant infection. Both the non-significance of a Shapiro-Wilk normality test (W = 0.99953, p-value = 0.4668) and visual inspection of a quantile-quantile plot indicate the our mixture model recapitulates the normal distribution anticipated for recipient log10 SpVL.

### Transmitter Allocation

The tolerance and heritability models were fitted to data pertaining to individuals from the Swiss HIV Cohort Study ^44,27^. 196 transmission pairs were previously inferred from monophyletic clusters of patients on maximum-likelihood phylogenetic trees ^43^. However, the direction of transmission within these pairs is unknown. To leverage these data in our model framework, we were required to impute an epidemiological role (transmitter/recipient) from the SpVLs. Initially, four allocation methods were trialled for SpVL, Vi, of each transmission pair, j,:

1. Random designation of transmitter

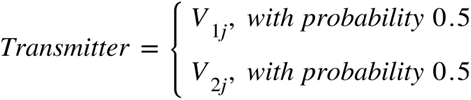

2. Greatest SpVL within a pair designated transmitter

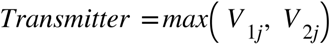

3. SpVL with highest likelihood of being associated(?) with a simulated distribution of transmitter SpVLs designated as transmitter

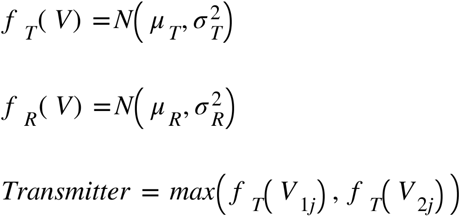

4. Epidemiological role simulated from the posterior probability of being a transmitter/recipient given simulated distributions of SpVL for transmitter and recipient.

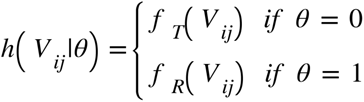

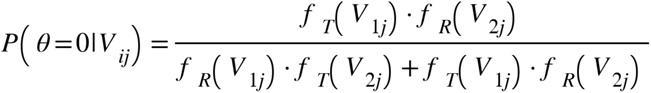

For methods 3 and 4, populations of transmitter and recipient SpVL were calculated from the linear equations described in the previous section. The initial carrier population was approximated by a Normal distribution with mean 4.35 and variance, 0.228 ^28,42^. When comparing the distributions of recipient and transmitter set point viral loads allocated by these four methods, we found that the maximum likelihood approach was consistent with repeated iterations of the Bayesian approach (Figure S2). The maximum likelihood approach was selected for the main analysis with random allocation maintained as a null scenario.

### Simulated Cohort Validation

To simulate risk group stratified cohorts, we first filtered the Swiss HIV Cohort transmission pairs to identify those with concordant risk groups. We then combined transmitter and recipient sex to form a ‘paired sex’ variable. We calculated the mean and variance-covariance matrices for each risk group of mean log10 SpVL, age at infection and paired sex. ‘Age at infection’ was log-transformed prior to simulation. These values parameterised a truncated multivariate normal distribution, from which combinations of values were drawn using rejection sampling.

Truncation was applied only to ‘age at infection’ with upper and lower bounds set at real numbers 16 and 80, respectively. The categorical variables of the new data set were set according to the cumulative probability distribution calculated for each variable from the empirical data. To validate these cohorts, we calculated the difference between the variances and normalised means of the simulated data from the empirical data across each risk group.

## Supplementary figures and tables

**Figure S1.**
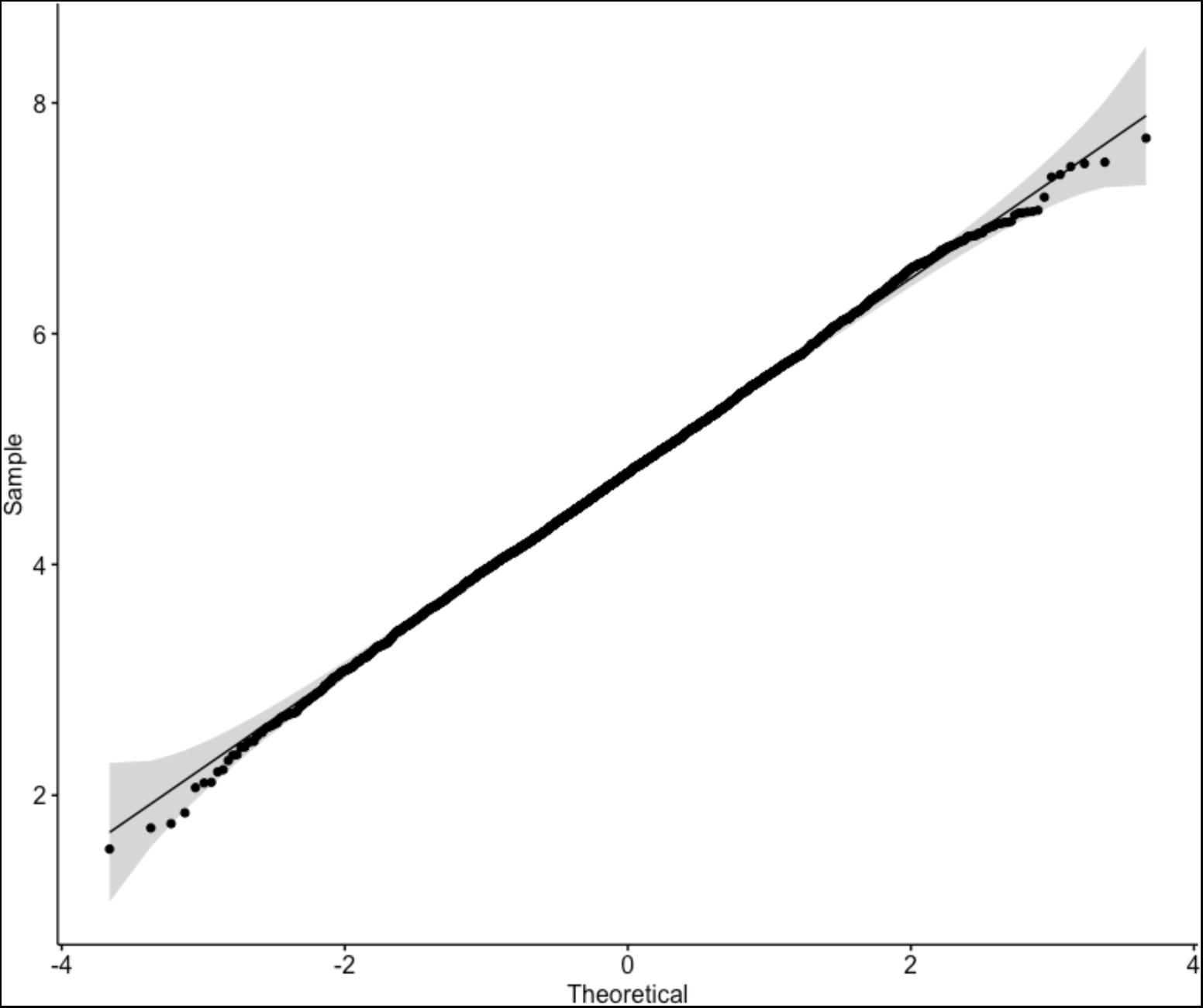
Quantile-Quantile plot of the combined recipient log10 SpVLs, sampled from multiple variant and single variant components of our Gaussian mixture model.

**Figure S2:**
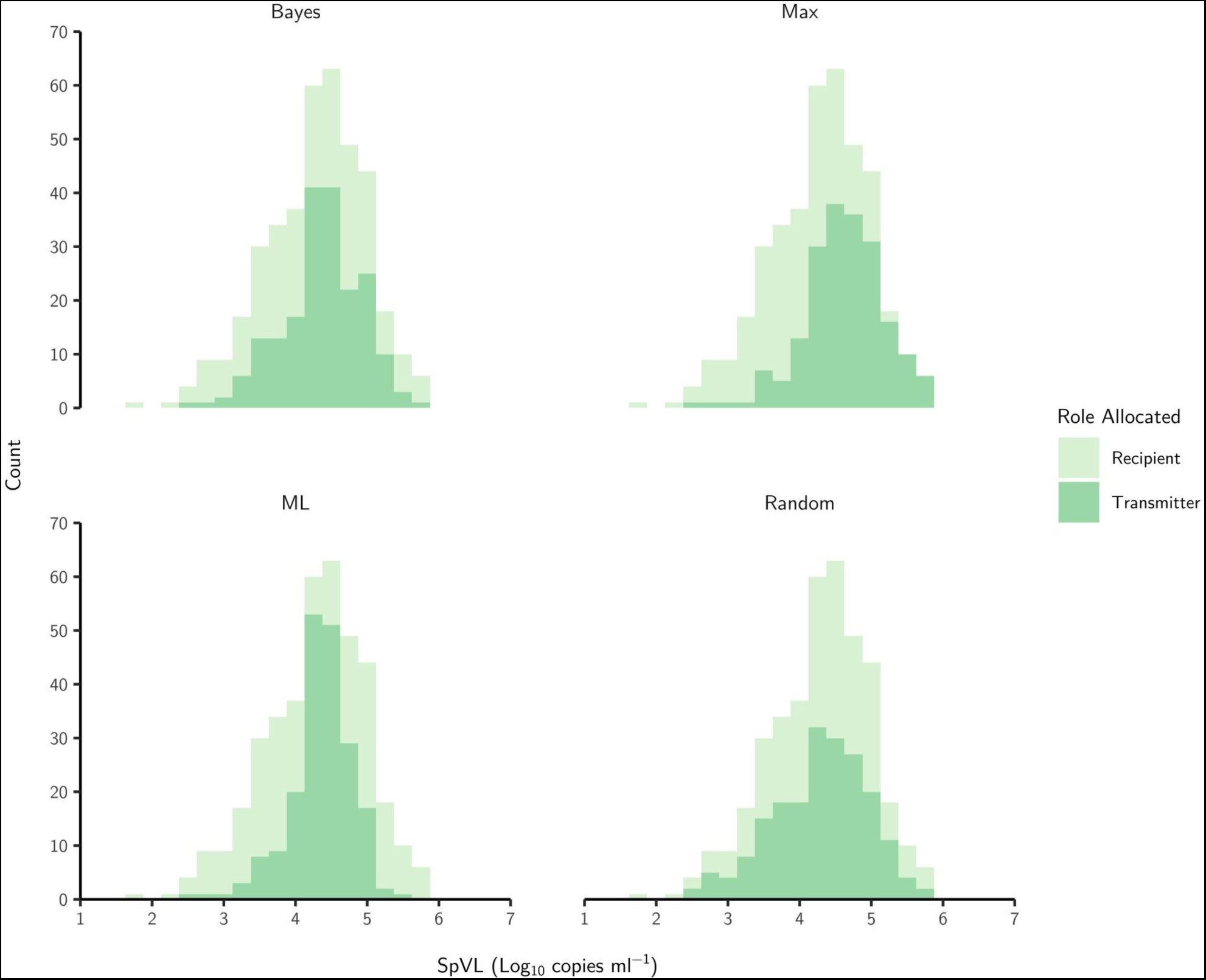
Histograms of log10 set point viral loads from 196 transmission pair of the Swiss HIV Cohort study. Transmitter/Recipient were allocated according to random, maximum SpVL, maximum likelihood (ML), and a posterior probability estimate (Bayes).

**Figure S3:**
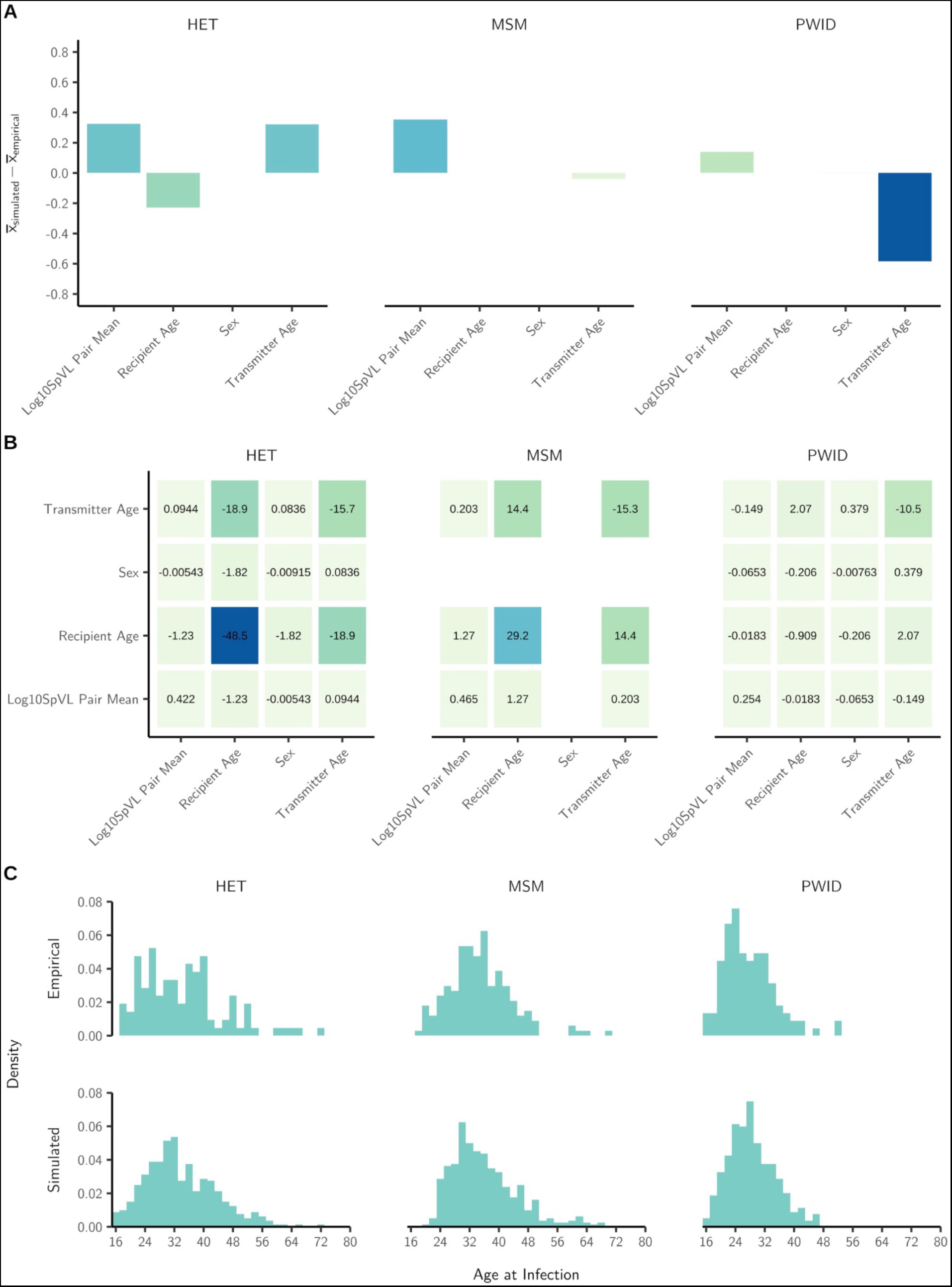
A) The difference of the variable means between simulated and empirical datasets. B) Differences of the variance-covariance matrices between simulated and empirical datasets. Darker hues correspond to a greater difference. C) Histograms of age at infection for simulated and empirical data, stratified by risk group.

**Figure S4:**
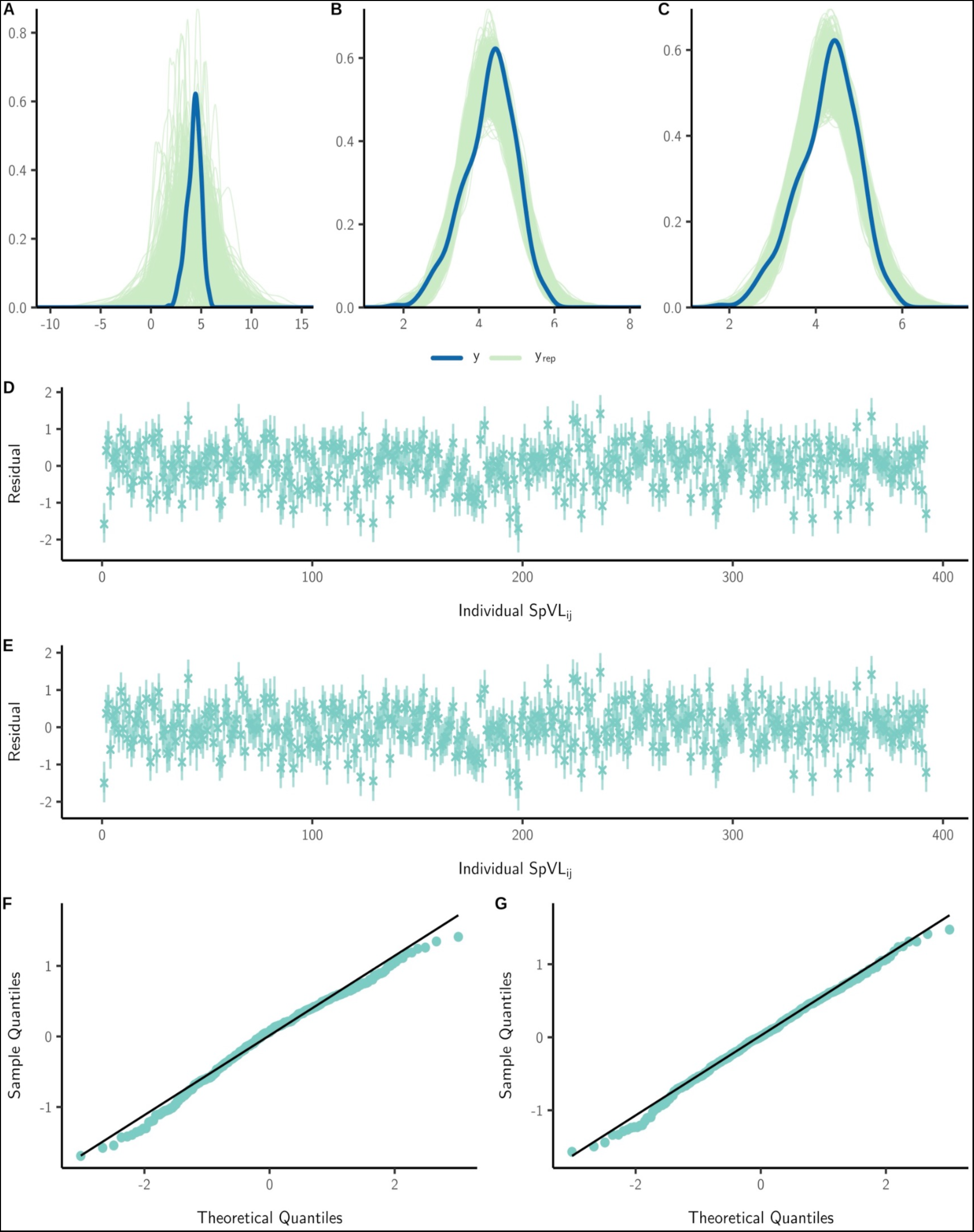
A) Comparing simulations from the prior-predictive distribution with observed values log10 SpVL to confirm our specified priors accurately reflected the empirical data. B+C) Simulations from the posterior-predictive distribution compared with observed values log10 SpVL indicate both models accurately recapitulate the empirical data. D+E) Median and 95% credible intervals summarising residual draws for each SpVL estimate in the random (D) and maximum likelihood (E) transmitter models. F+G) Linearity of points on quantile-quantile plots for both random (F) and maximum likelihood (G) allocated transmitter models suggest these data followed a Normal distribution.

**Figure S5:**
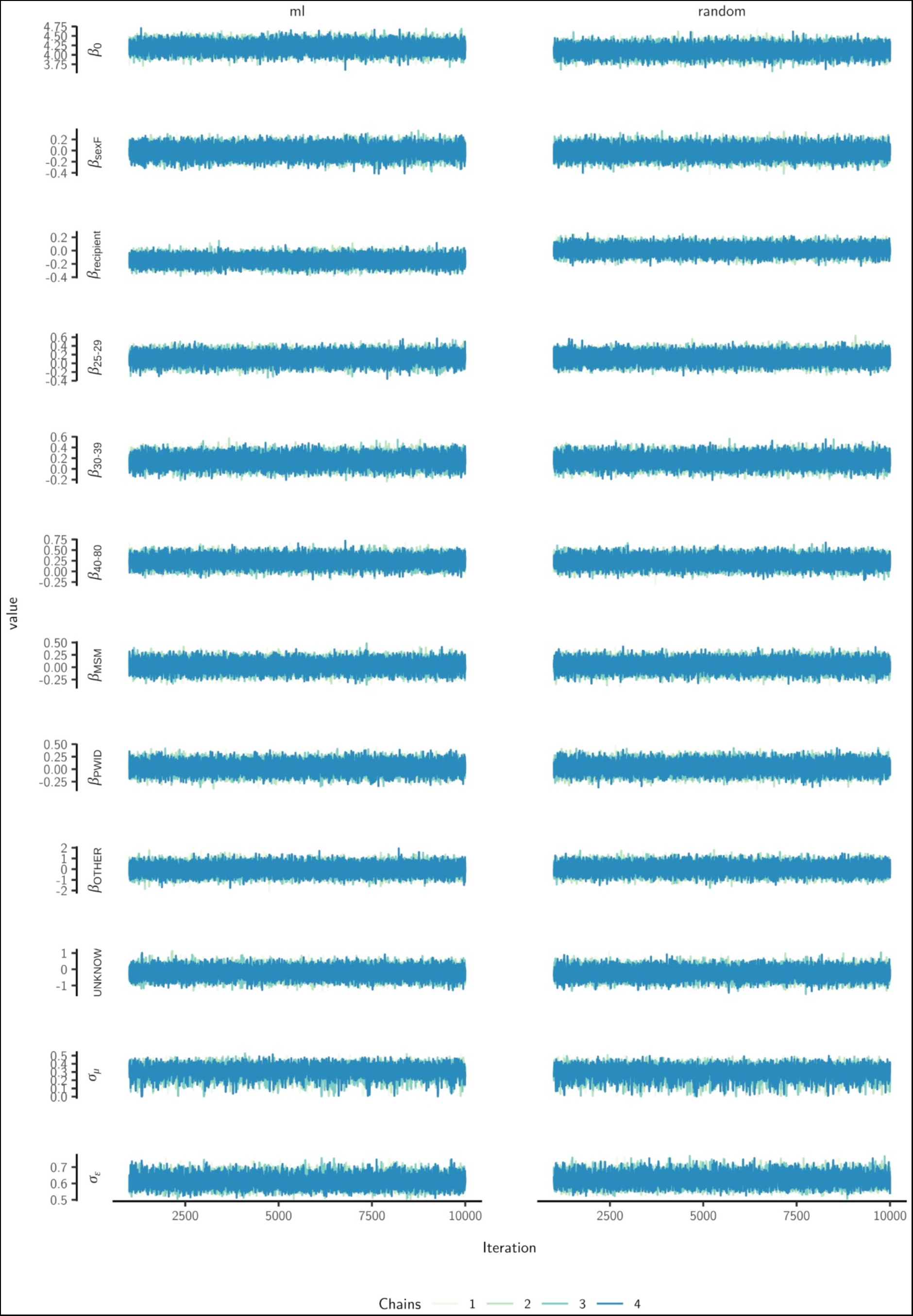
MCMC traces (post burn-in) for each parameter value fitted in the heritability model.

**Figure S6:**
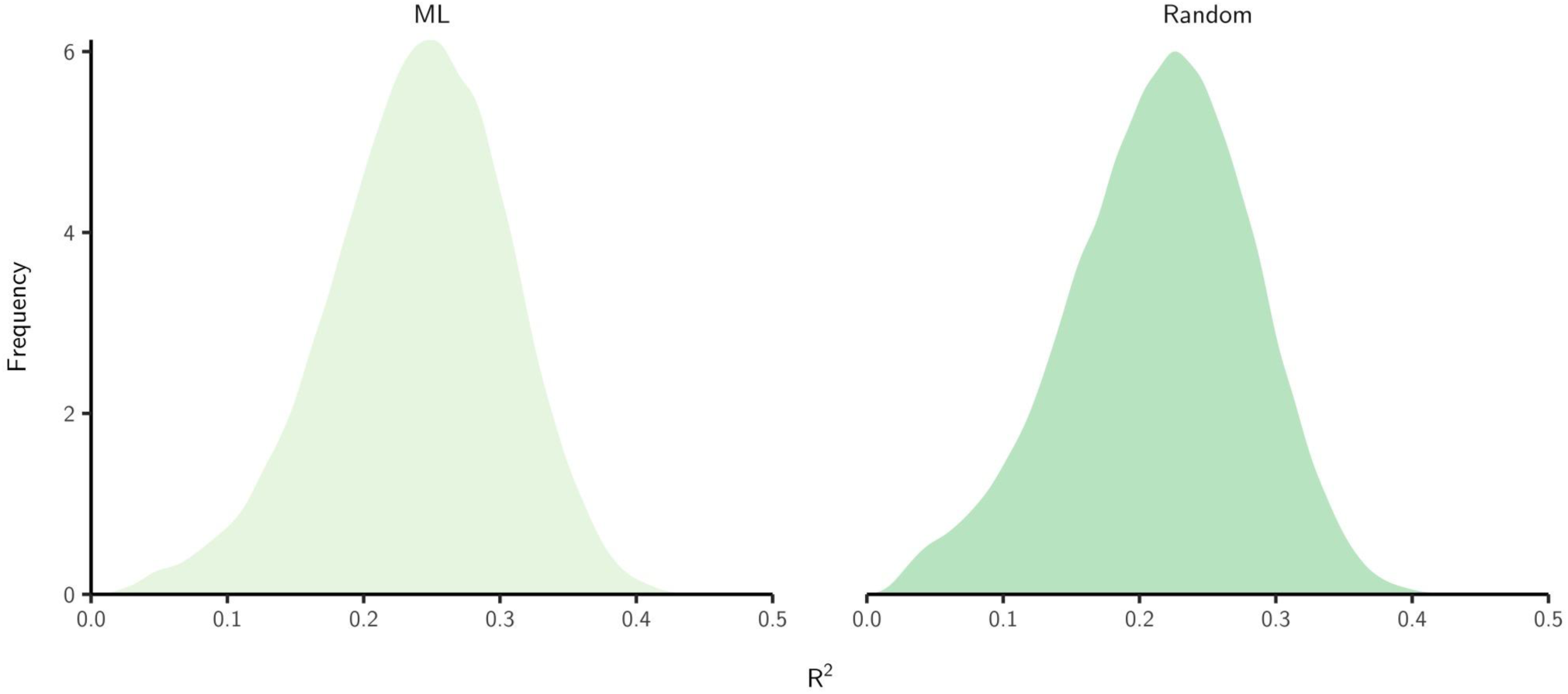
Posterior distributions of our estimate of broad-sense heritability, H^2^; approximated by R^2^ estimated using a reformulation described by Gelman et al[9]. For the random transmitter allocation model, R^2^ was 0.224 (95% Highest Posterior Density (HPD): 0.110-0.326), and increased to 0.239 (0.123-0.341) for the maximum likelihood allocated transmitter model.

**Figure S7:**
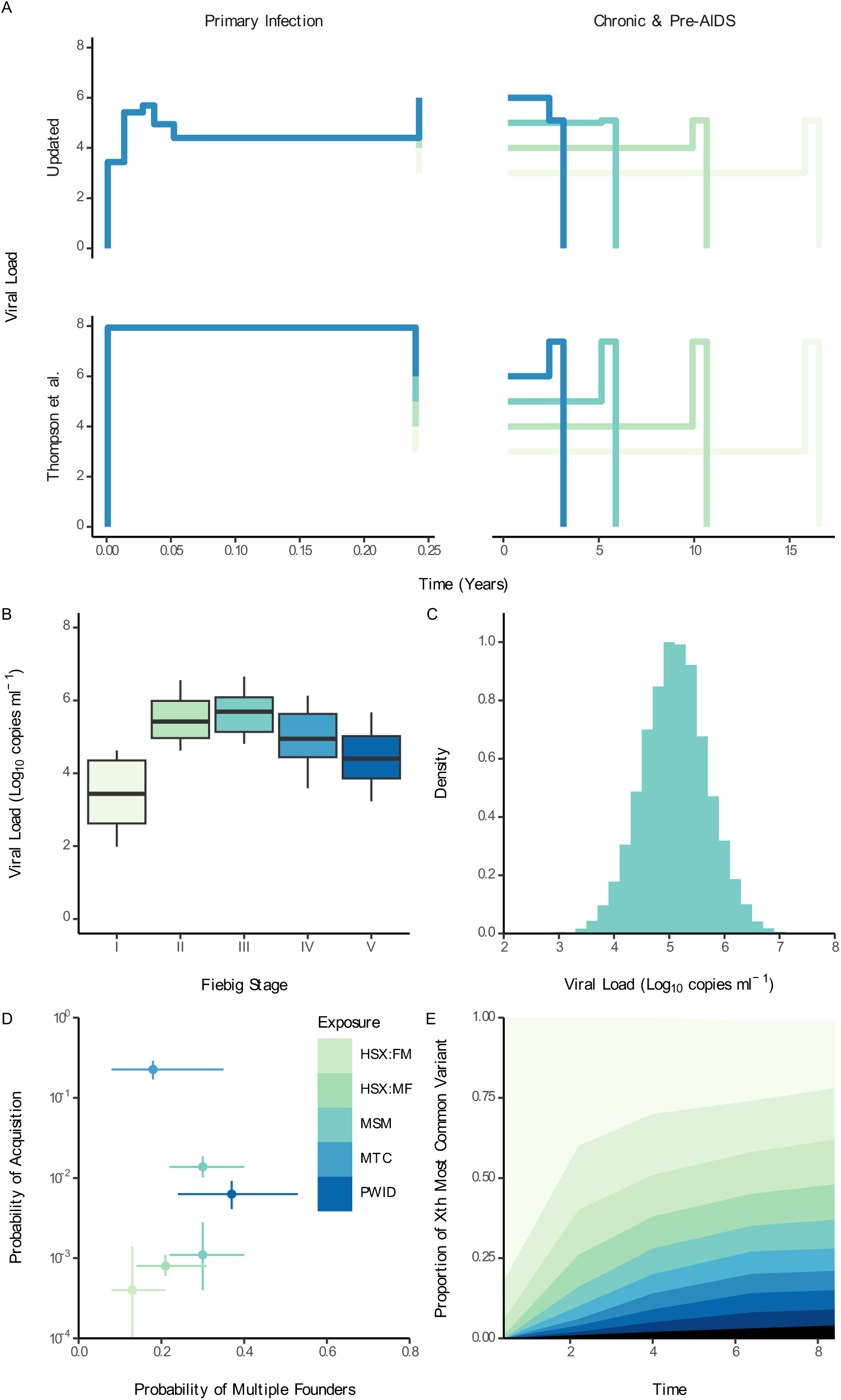
Key components of the transmission model. A) The per-event transmission probability is the zero-inflated density function of the number of transmitted virions and variants, sampled from the total viral load. Our updated model leverages data from Fiebig et al. (B) and Mellors et al. (C) to better define acute and pre-aids viral loads respectively ^48,49^. Our updated model is then fitted to previously published estimates of the per-event probability of acquisition and the probability that infection is initiated by multiple founder variants (D) ^19,51^. The model describing within-patient viral evolution remains unchanged from Thompson et al (E) ^26,50^

**Figure S8:**
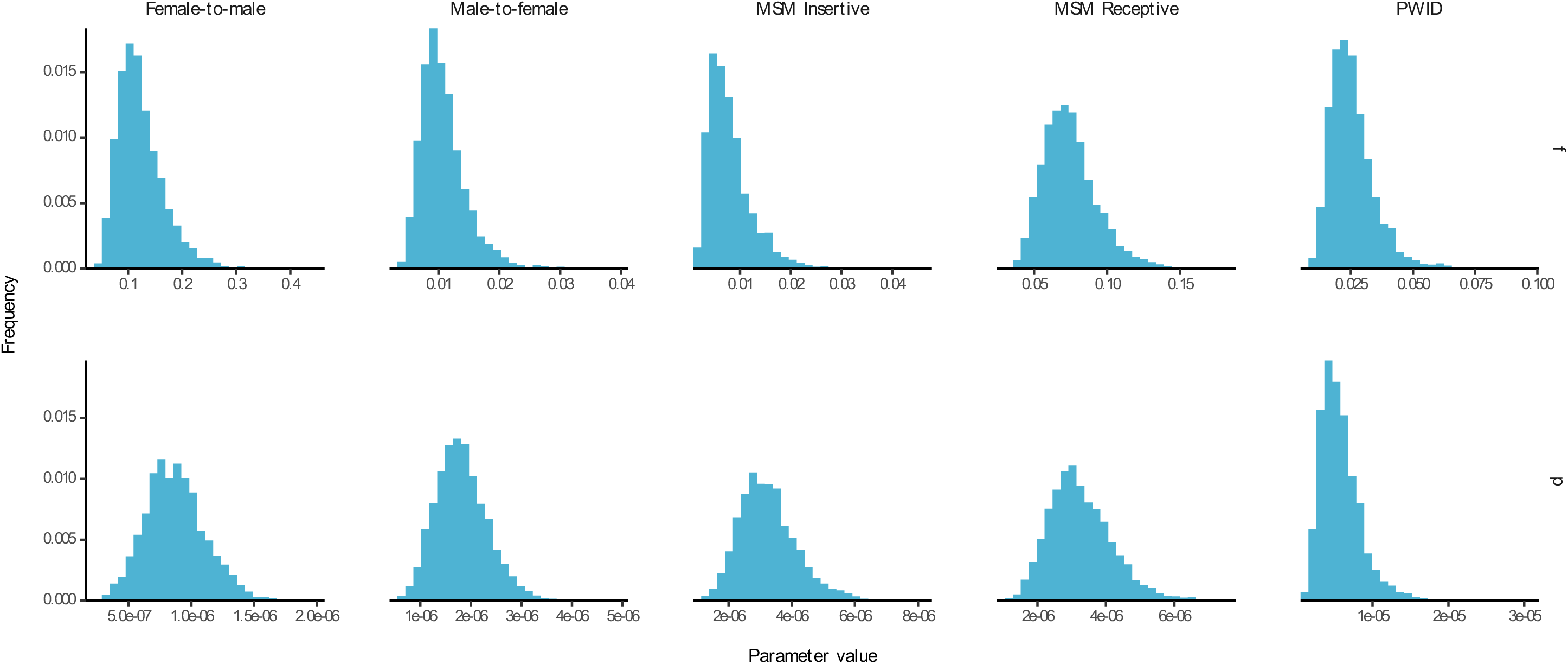
Joint posterior distributions of the zero-inflation parameter f (the proportion of environments in which transmission is possible) and per-particle probability of transmission, p, as estimated in our updated transmission model.

**Figure S9:**
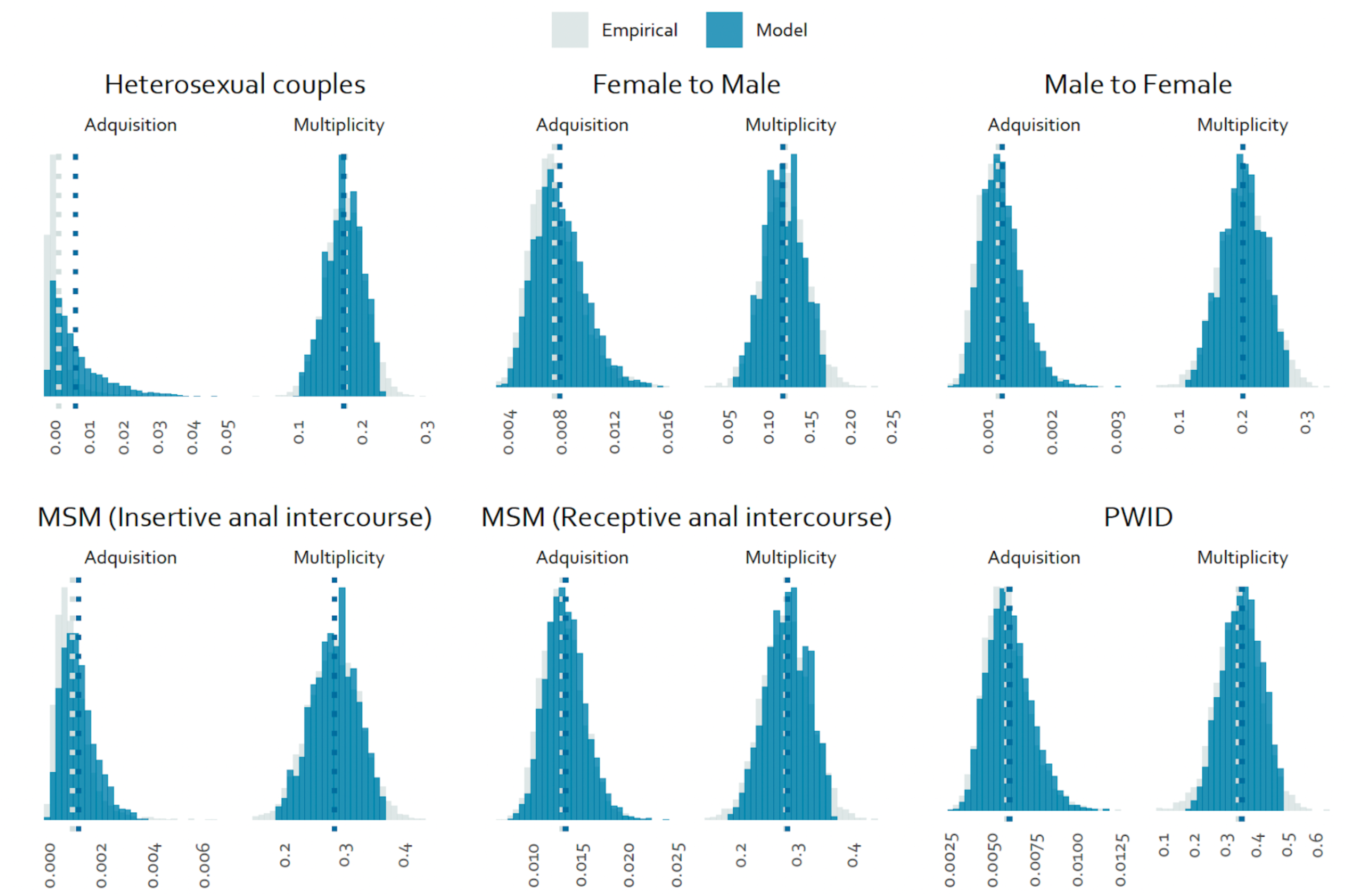
Joint posterior distributions of the probability that infection is initiated by multiple variants and the probability of infection acquisition, superimposed over previous empirical characterisations^51,19^

**Figure S10:**
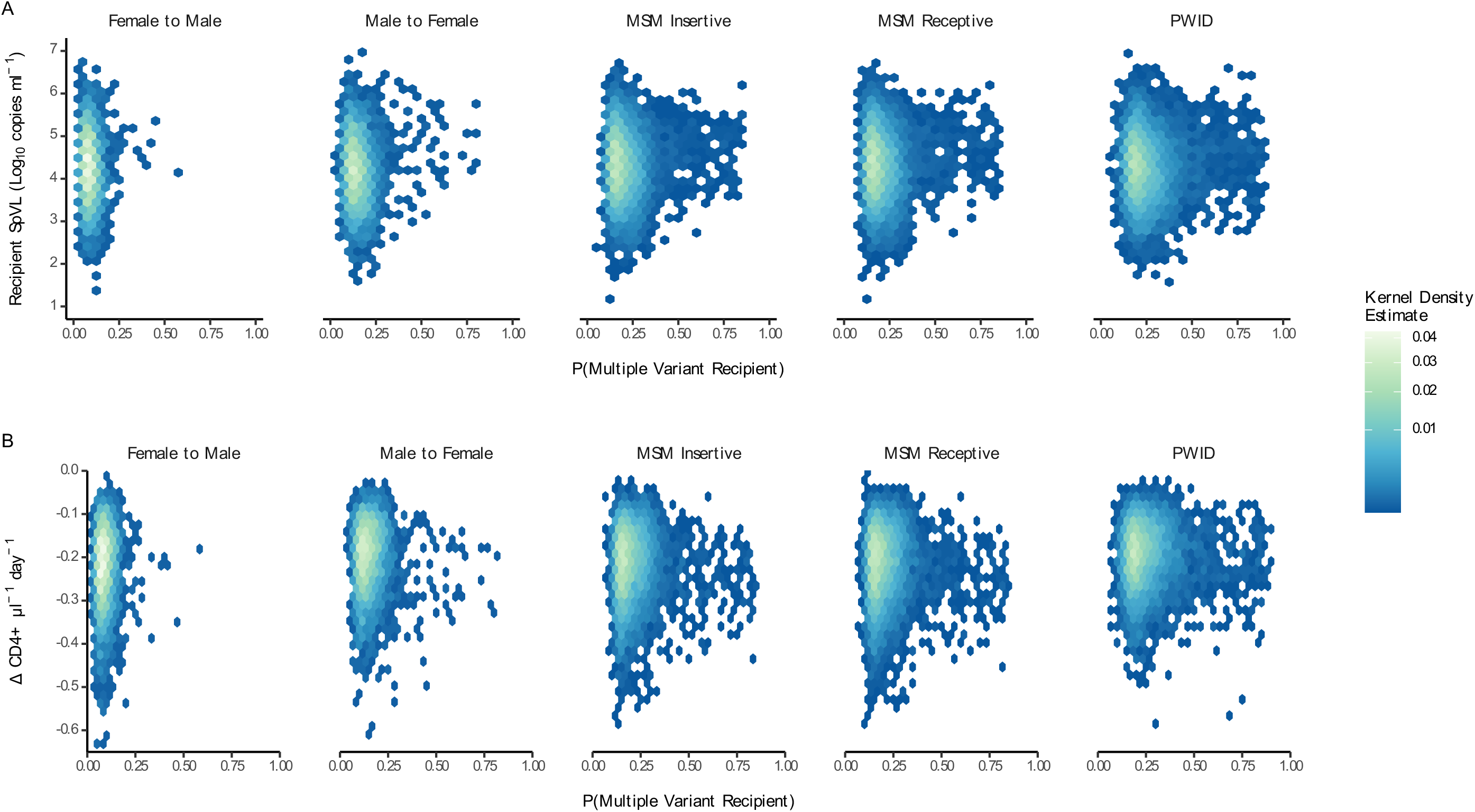
The relationship between the probability that infection is initiated by multiple variants intermediate, SpVL (A) with CD4+ T cell decline (B) was consistent across transmitter allocation. In our main analysis we imputed transmitter status by maximum likelihood. Here, transmitter status was assigned to the individual within the transmission pair with the greatest viral load.

**Figure S11:**
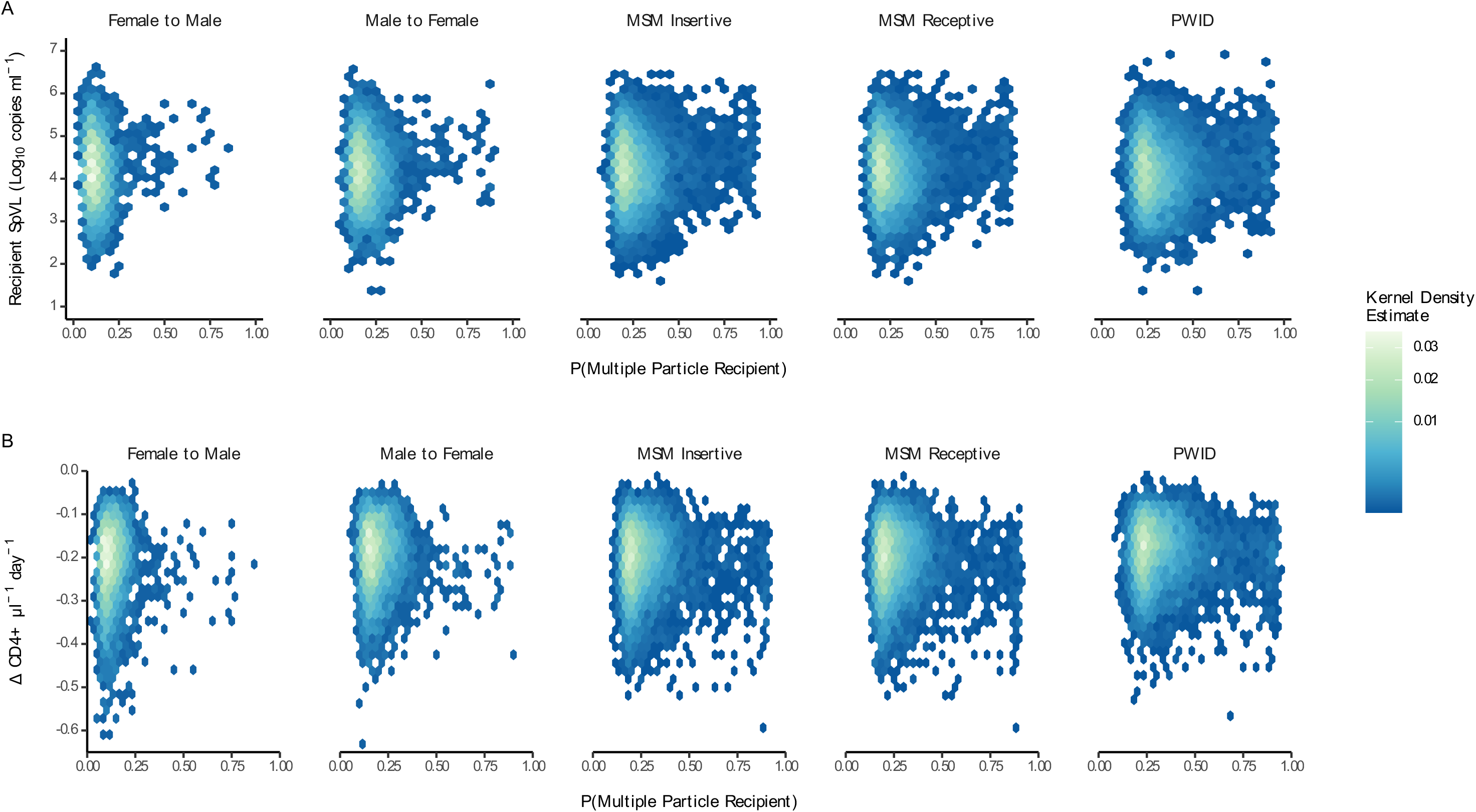
The relationship between the probability that infection is initiated by multiple viral particles was also consistent with our main analysis, showing that recipient infections were most likely to have a low probability of being initiated by multiple variants and present with intermediate viral loads. (A) with sign of increased rate of CD4+ T cell decline (B).

**Table S1:**
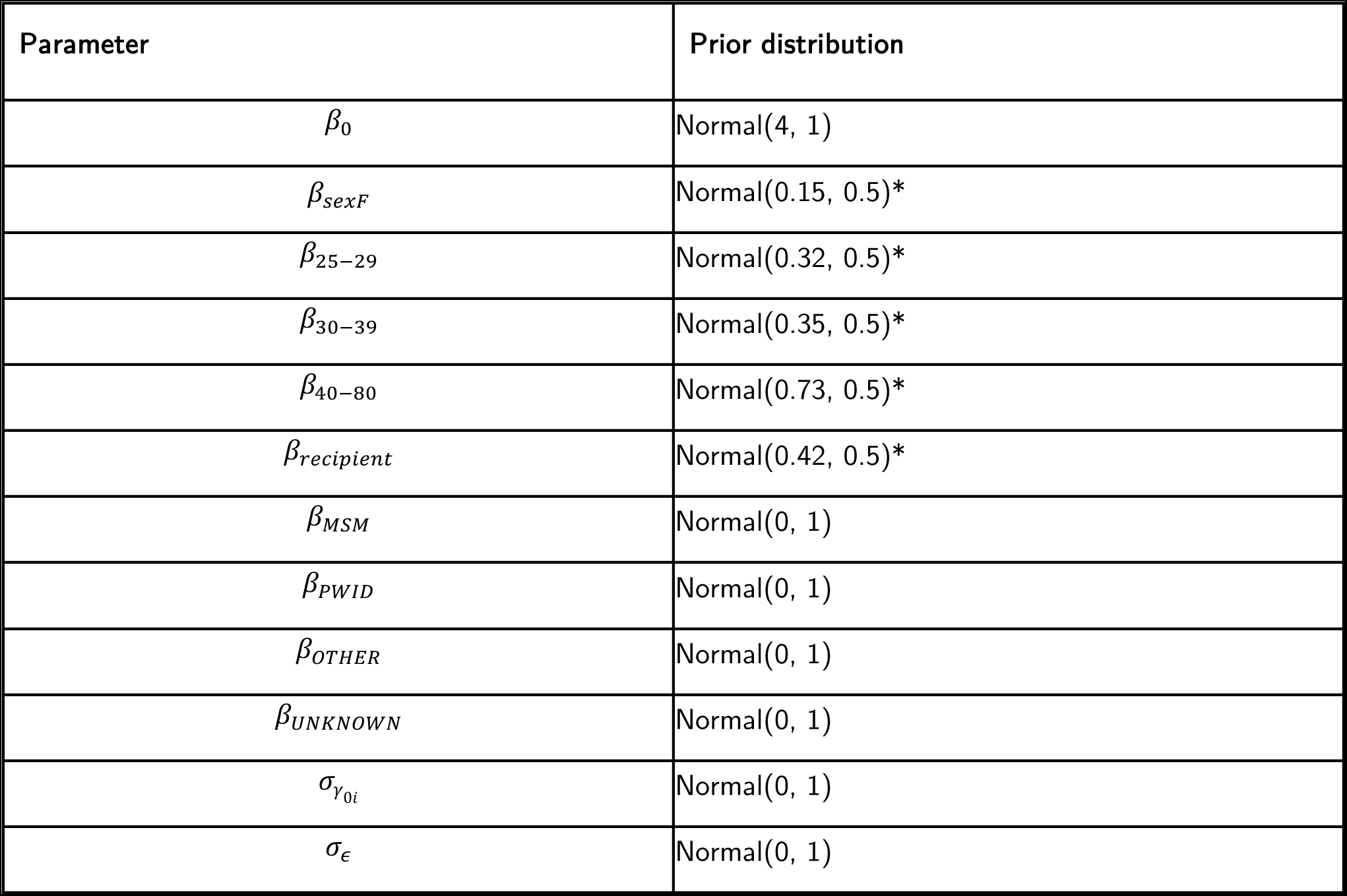
Priors used for Bayesian linear mixed model to estimate heritability of SpVL. *= Priors informed by the analysis of Hollingsworth et al. ^21^

| Parameter | Prior distribution |
| --- | --- |
| $\beta_0$ | Normal(4, 1) |
| $\beta_{sexF}$ | Normal(0.15, 0.5)* |
| $\beta_{25-29}$ | Normal(0.32, 0.5)* |
| $\beta_{30-39}$ | Normal(0.35, 0.5)* |
| $\beta_{40-80}$ | Normal(0.73, 0.5)* |
| $\beta_{recipient}$ | Normal(0.42, 0.5)* |
| $\beta_{MSM}$ | Normal(0, 1) |
| $\beta_{PWID}$ | Normal(0, 1) |
| $\beta_{OTHER}$ | Normal(0, 1) |
| $\beta_{UNKNOWN}$ | Normal(0, 1) |
| $\sigma_{\gamma_{0i}}$ | Normal(0, 1) |
| $\sigma_{\epsilon}$ | Normal(0, 1) |

**Table S2:** Effective sample sizes (ESS) and Gelman-Rubin values (X=1 implies convergence) for each parameter value fitted in the heritability model.

| Parameter Value | Transmitter Allocation: Random |  | Transmitter Allocation: ML SpVL |  |
| --- | --- | --- | --- | --- |
|  | ESS | Gelman-Rubin Diagnostic | ESS | Gelman-Rubin Diagnostic |
| $\beta_0$ | 19142 | 1.00 | 28739 | 1.00 |
| $\beta_{sexF}$ | 29647 | 1.00 | 44591 | 1.00 |
| $\beta_{25-29}$ | 28118 | 1.00 | 35379 | 1.00 |
| $\beta_{30-39}$ | 24191 | 1.00 | 31774 | 1.00 |
| $\beta_{40-80}$ | 24128 | 1.00 | 34721 | 1.00 |
| $\beta_{recipient}$ | 48955 | 1.00 | 66927 | 1.00 |
| $\beta_{MSM}$ | 23699 | 1.00 | 35896 | 1.00 |
| $\beta_{PWID}$ | 21533 | 1.00 | 30231 | 1.00 |
| $\beta_{OTHER}$ | 39539 | 1.00 | 59339 | 1.00 |
| $\beta_{UNKNOWN}$ | 38137 | 1.00 | 47706 | 1.00 |
| $\sigma_{\gamma_{0i}}$ | 4967 | 1.00 | 11437 | 1.00 |
| $\sigma_{\epsilon}$ | 7599 | 1.00 | 15192 | 1.00 |

**Table S3:**
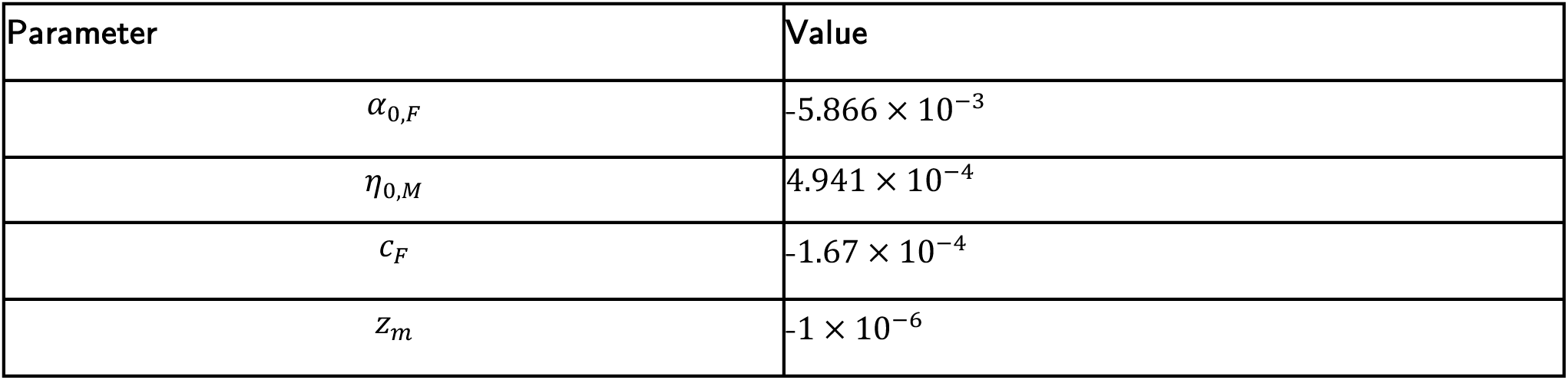
Fixed parameter values used for the tolerance model. Parameter values extracted from Regoes et al. ^27^

**Table S4.** Fixed parameter values used for the tolerance model. Parameter values extracted from Thompson et al. ^26^

| Parameter | Value |
| --- | --- |
| $\alpha$ | -3.55 |
| Dmax | 25.4 |
| D50 | 3058 |
| Dk | 0.41 |
| taup | 0.24 |
| tauc | 0.75 |

## References

1. Isaksson, B. et al. AIDS Two Months After Primary Human Immunodeficiency Virus Infection. The Journal of Infectious Diseases 158, 866–868 (1988).

2. Buchbinder, S. P., Katz, M. H., Hessol, N. A., O’Malley, P. M. & Holmberg, S. D. Long-term HIV-1 infection without immunologic progression. AIDS 8, 1123 (1994).

3. Mellors, J. W. et al. Prognosis in HIV-1 Infection Predicted by the Quantity of Virus in Plasma. Science 272, 1167 LP – 1170 (1996).

4. Bartha, I., Simon, P. & Müller, V. Has HIV evolved to induce immune pathogenesis? Trends in Immunology 29, 322–328 (2008).

5. Carlson, J. M. et al. Impact of pre-adapted HIV transmission. Nat Med 22, 606–613 (2016).

6. Rosenberg, P. S., Goedert, J. J. & Biggar, R. J. Effect of age at seroconversion on the natural AIDS incubation distribution. Multicenter Hemophilia Cohort Study and the International Registry of Seroconverters. AIDS 8, 803–810 (1994).

7. Fellay, J. et al. Common genetic variation and the control of HIV-1 in humans. PLoS genetics 5, e1000791 (2009).

8. Bhattacharya, T. et al. Founder Effects in the Assessment of HIV Polymorphisms and HLA Allele Associations. Science 315, 1583–1586 (2007).

9. Best, K., Barouch, D. H., Guedj, J., Ribeiro, R. M. & Perelson, A. S. Zika virus dynamics: Effects of inoculum dose, the innate immune response and viral interference. PLOS Computational Biology 17, e1008564 (2021).

10. Han, A. et al. A Dose-finding Study of a Wild-type Influenza A(H3N2) Virus in a Healthy Volunteer Human Challenge Model. Clinical Infectious Diseases 69, 2082–2090 (2019).

11. Sagar, M. et al. Infection with Multiple Human Immunodeficiency Virus Type 1 Variants Is Associated with Faster Disease Progression. Journal of Virology 77, 12921–12926 (2003).

12. Abrahams, M.-R. et al. Quantitating the Multiplicity of Infection with Human Immunodeficiency Virus Type 1 Subtype C Reveals a Non-Poisson Distribution of Transmitted Variants. Journal of Virology 83, 3556–3567 (2009).

13. Janes, H. et al. HIV-1 infections with multiple founders are associated with higher viral loads than infections with single founders. Nat Med 21, 1139–1141 (2015).

14. Chaillon, A. et al. Characterizing the multiplicity of HIV founder variants during sexual transmission among MSM. Virus Evolution 2, (2016).

15. Macharia, G. N. et al. Infection with multiple HIV-1 founder variants is associated with lower viral replicative capacity, faster CD4+ T cell decline and increased immune activation during acute infection. PLoS pathogens 16, e1008853–e1008853 (2020).

16. Zhu, T. et al. Genotypic and phenotypic characterization of HIV-1 patients with primary infection. Science 261, 1179 LP – 1181 (1993).

17. Zhang, L. et al. Selection for specific sequences in the external envelope protein of human immunodeficiency virus type 1 upon primary infection. Journal of Virology 67, 3345–3356 (1993).

18. Keele, B. F. et al. Identification and characterization of transmitted and early founder virus envelopes in primary HIV-1 infection. Proceedings of the National Academy of Sciences 105, 7552–7557 (2008).

19. Baxter, J. et al. Inferring the multiplicity of founder variants initiating HIV-1 infection: a systematic review and individual patient data meta-analysis. The Lancet Microbe (2023) doi:10.1016/S2666-5247(22)00327-5.

20. Song, H. et al. Transmission of Multiple HIV-1 Subtype C Transmitted/founder Viruses into the Same Recipients Was not Determined by Modest Phenotypic Differences. Scientific Reports 6, 38130 (2016).

21. Hollingsworth, T. D. et al. HIV-1 Transmitting Couples Have Similar Viral Load Set-Points in Rakai, Uganda. PLOS Pathogens 6, e1000876 (2010).

22. van der Kuyl, A. C., Jurriaans, S., Pollakis, G., Bakker, M. & Cornelissen, M. HIV RNA levels in transmission sources only weakly predict plasma viral load in recipients. AIDS 24, 1607 (2010).

23. Alizon, S. et al. Phylogenetic Approach Reveals That Virus Genotype Largely Determines HIV Set-Point Viral Load. PLOS Pathogens 6, e1001123 (2010).

24. Blanquart, F. et al. Viral genetic variation accounts for a third of variability in HIV-1 set-point viral load in Europe. PLoS Biol 15, e2001855–e2001855 (2017).

25. Hodcroft, E. et al. The Contribution of Viral Genotype to Plasma Viral Set-Point in HIV Infection. PLOS Pathogens 10, e1004112 (2014).

26. Thompson, R. N. et al. Link between the numbers of particles and variants founding new HIV-1 infections depends on the timing of transmission. Virus Evolution 5, (2019).

27. Regoes, R. R. et al. Disentangling Human Tolerance and Resistance Against HIV. PLOS Biology 12, e1001951 (2014).

28. Fraser, C., Hollingsworth, T. D., Chapman, R., de Wolf, F. & Hanage, W. P. Variation in HIV-1 set-point viral load: Epidemiological analysis and an evolutionary hypothesis. Proceedings of the National Academy of Sciences 104, 17441–17446 (2007).

29. Haaland, R. E. et al. Inflammatory Genital Infections Mitigate a Severe Genetic Bottleneck in Heterosexual Transmission of Subtype A and C HIV-1. PLOS Pathogens 5, e1000274 (2009).

30. Carlson, J. M. et al. Selection bias at the heterosexual HIV-1 transmission bottleneck. Science 345, 1254031 (2014).

31. Kijak, G. H. et al. Rare HIV-1 transmitted/founder lineages identified by deep viral sequencing contribute to rapid shifts in dominant quasispecies during acute and early infection. PLoS pathogens 13, e1006510–e1006510 (2017).

32. Fischer, W. et al. Transmission of single HIV-1 genomes and dynamics of early immune escape revealed by ultra-deep sequencing. PloS one 5, e12303–e12303 (2010).

33. Li, H. et al. High Multiplicity Infection by HIV-1 in Men Who Have Sex with Men. PLOS Pathogens 6, e1000890 (2010).

34. Song, H. et al. Tracking HIV-1 recombination to resolve its contribution to HIV-1 evolution in natural infection. Nature communications 9, 1928 (2018).

35. Lewitus, E. et al. HIV-1 infections with multiple founders associate with the development of neutralization breadth. PLOS Pathogens 18, e1010369 (2022).

36. Joyce, C. et al. Antigen pressure from two founder viruses induces multiple insertions at a single antibody position to generate broadly neutralizing HIV antibodies. PLOS Pathogens 19, e1011416 (2023).

37. Altfeld, M. et al. Influence of HLA-B57 on clinical presentation and viral control during acute HIV-1 infection. AIDS 17, 2581 (2003).

38. Daar, E. S. et al. Baseline HIV Type 1 Coreceptor Tropism Predicts Disease Progression. Clinical Infectious Diseases 45, 643–649 (2007).

39. Mitov, V. & Stadler, T. A Practical Guide to Estimating the Heritability of Pathogen Traits. Molecular Biology and Evolution 35, 756–772 (2018).

40. Bertels, F. et al. Dissecting HIV Virulence: Heritability of Setpoint Viral Load, CD4+ T-Cell Decline, and Per-Parasite Pathogenicity. Molecular Biology and Evolution 35, 27–37 (2018).

41. Bachmann, N. et al. Parent-offspring regression to estimate the heritability of an HIV-1 trait in a realistic setup. Retrovirology 14, 33 (2017).

42. Bonhoeffer, S., Fraser, C. & Leventhal, G. E. High Heritability Is Compatible with the Broad Distribution of Set Point Viral Load in HIV Carriers. PLOS Pathogens 11, e1004634 (2015).

43. Kouyos, R. D. et al. Clustering of HCV coinfections on HIV phylogeny indicates domestic and sexual transmission of HCV. International Journal of Epidemiology 43, 887–896 (2014).

44. Scherrer, A. U. et al. Cohort Profile Update: The Swiss HIV Cohort Study (SHCS). International Journal of Epidemiology 51, 33–34j (2022).

45. Vehtari, A., Gelman, A., Simpson, D., Carpenter, B. & Bürkner, P.-C. Rank-Normalization, Folding, and Localization: An Improved R^ for Assessing Convergence of MCMC (with Discussion). Bayesian Analysis 16, 667–718 (2021).

46. Gelman, A., Goodrich, B., Gabry, J. & Vehtari, A. R-squared for Bayesian Regression Models. The American Statistician 73, 307–309 (2019).

47. Rodríguez, B. et al. Predictive Value of Plasma HIV RNA Level on Rate of CD4 T-Cell Decline in Untreated HIV Infection. JAMA 296, 1498–1506 (2006).

48. Fiebig, E. W. et al. Dynamics of HIV viremia and antibody seroconversion in plasma donors: implications for diagnosis and staging of primary HIV infection. AIDS 17, 1871 (2003).

49. Mellors, J. W. et al. Quantitation of HIV-1 RNA in Plasma Predicts Outcome after Seroconversion. Ann Intern Med 122, 573–579 (1995).

50. Zanini, F. et al. Population genomics of intrapatient HIV-1 evolution. eLife 4, e11282 (2015).

51. Patel, P. et al. Estimating per-act HIV transmission risk: a systematic review. AIDS 28, 1509 (2014).

52. R Core Team. R: A Language and Environment for Statistical Computing. (R Foundation for Statistical Computing, Vienna, Austria., 2018).

53. Carpenter, B. et al. Stan: A Probabilistic Programming Language. Journal of Statistical Software 76, 1–32 (2017).

54. Bürkner, P.-C. brms: An R Package for Bayesian Multilevel Models Using Stan. Journal of Statistical Software 80, 1–28 (2017).

55. Gabry, J., Simpson, D., Vehtari, A., Betancourt, M. & Gelman, A. Visualization in Bayesian Workflow. Journal of the Royal Statistical Society Series A: Statistics in Society 182, 389–402 (2019).

56. Wickham, H. et al. Welcome to the Tidyverse. Journal of open source software 4, 1686 (2019).

57. Kay, M. tidybayes: Tidy Data and Geoms for Bayesian Models. R package version 2.1. 1. (2020).

58. Lüdecke, D., Ben-Shachar, M. S., Patil, I., Waggoner, P. & Makowski, D. performance: An R package for assessment, comparison and testing of statistical models. Journal of Open Source Software 6, (2021).

